# Brain heterogeneity in 1,792 individuals with schizophrenia: effects of illness stage, sites of origin and pathophysiology

**DOI:** 10.1101/2024.05.23.24307840

**Authors:** Yuchao Jiang, Lena Palaniyappan, Xiao Chang, Jie Zhang, Enpeng Zhou, Xin Yu, Shih-Jen Tsai, Ching-Po Lin, Jingliang Cheng, Yingying Tang, Jijun Wang, Cheng Luo, Dezhong Yao, Long-Biao Cui, Wei Cheng, Jianfeng Feng

## Abstract

**Importance:** Schizophrenia is characterized with greater variability beyond the mean differences in brain structures. This variability is often assumed to be static, reflecting the presence of heterogeneous subgroups, but this assumption and alternative explanations remain untested.

**Objective:** To test if gray matter volume (GMV) variability is more less in later stages of schizophrenia, and evaluate if a putative ‘spreading pattern’ with GMV deficits originating in one part of the brain and diffusing elsewhere explain the variability of schizophrenia.

**Design, settings, and participants:** This study evaluated the regional GMV variability using MRI of 1,792 individuals with schizophrenia and 1,523 healthy controls (HCs), and the association of GMV variability with neurotransmitter and transcriptomic gene data in the human brain.

**Main outcomes and measures:** Regional variability was evaluated by comparing the relative variability of patients to controls, using the relative mean-scaled log variability ratio (lnCVR). A network diffusion model (NDM) was employed to simulate the possible processes of GMV alteration across brain regions.

**Results:** Compared with HCs, greater lnCVR (*p_FDR_*<0.05) was found in 50 regions in the whole patient group (n=1792; 762 females; mean[SD] age, 29.9[11.9] years), at a much greater frequency (*p=5.0*×10^−13^) in the first-episode drug-naïve subsample (73 regions) (n=478; mean[SD] illness duration, 0.548[0.459] years), compared to the chronic medicated subsample (28 regions) (n=398; mean[SD] illness duration, 14.0[10.4] years). The average lnCVR across all regions was greater in the first-episode than chronic subsample (*t*=10.8, *p=*1.7×10^−7^). The areas with largest lnCVR were located at frontotemporal cortex and thalamus (first-episode), or hippocampus and caudate (chronic); there was a significant correlation with case-control mean difference (*r*=0.367, *p=*6.7×10^−4^). We determined a gene expression map that correlated with the lnCVR map in schizophrenia (*r*=0.491, *p*=0.003). The NDM performed consistently (72.1% patients, *p_spin_*<0.001) in replicating GMV changes when simulated and observed values were compared.

**Conclusion and relevance:** Brain-based heterogeneity is unlikely to be a static feature of schizophrenia; it is more pronounced at the onset of the disorder but reduced over the long term. Differences in the site of ‘origin’ of GMV changes in individual-level may explain the observed anatomical variability in schizophrenia.

**Key Points:** *Question:* No two individuals with schizophrenia have the same anatomical change in the brain. Is this variability a fixed feature of schizophrenia or does it become more pronounced at later stages? Is this variability explained by a putative ‘spreading pattern’ of gray matter deficits originating in one part of the brain and diffusing elsewhere?

*Findings:* In 1,792 individuals with schizophrenia, neuroanatomical variability is not a fixed feature; it is more pronounced at the illness onset but less prominent in later stages. The neuroanatomical variability is associated with various molecular and neurobiological processes implicated in the neurodevelopmental etiology of schizophrenia. Differences in the site of ‘origin’ of gray matter deficits in each individual with schizophrenia explains most of the observed variability.

*Meaning:* Our work finds support for a space-time interaction along a shared pathophysiological continuum (network-based trans-neuronal diffusion), as a possible explanatory model for inter-subject variability. These findings contribute to the understanding that inter-individual variability in schizophrenia may arise from a common cohesive process that varies in its state (across time) and space (across brain regions). This also raises the question of what dynamic processes contribute to the reducing heterogeneity over time in schizophrenia. Answering this question will be a key test to the neurobiological validity of the concept of schizophrenia.

## 1. Introduction

Many psychiatric disorders, including schizophrenia, are heterogeneous in their clinical presentation and phenotypic markers such as brain measures. This heterogeneity may result from the multitude of mechanisms that contribute to the eventual clinical features, resulting from a lack of single genetic, molecular, cellular or mesoscale brain markers of schizophrenia [1, 2]. On the other hand, this also provides an opportunity for precision medicine approaches. For example, we may be able to ‘split’ schizophrenia into biologically more homogeneous subtypes [3], identified from phenotypic differences (e.g. brain structure [4]), that may differ in their clinical trajectories and respond differently to treatments [5, 6]. As a result, phenotypic heterogeneity is increasingly being viewed as a feature of interest in the causal models of schizophrenia.

One of the expectations when studying brain-based heterogeneity in schizophrenia is that the homogenously affected regions reflect a common pathophysiological process, while regions with high variability reflect non-overlapping subtypes [1, 7]. This expectation follows an implicit assumption that regional heterogeneity is a static feature that does not vary over illness course, which is yet to be tested in schizophrenia. Secondly, if high variability is concentrated in regions without a notable mean difference (i.e., no schizophrenia vs. controls effect), this supports the existence of different subtypes with no overlapping structural effects. On the other hand, the regions that show highest mean difference may also show the highest variability, if the same causal mechanism results in quantitative variations due to compensatory forces operating at these regions [8, 9]. The relationship between mean effect and variability is yet to be studied in schizophrenia.

Structural brain imaging studies consistently demonstrate lower gray matter volume (GMV) in schizophrenia [10, 11], which progresses over time across various stages of the illness [12–14]. Interestingly, recent studies suggest that the spatial pattern of GMV alterations associated with schizophrenia is not random, but rather shaped by the underlying architecture of brain networks [15]. In the context of anatomical heterogeneity, this opens up the possibility that each person with schizophrenia may have a distinct site of origin of GMV changes, but a diffusion or transneuronal ‘spread’ of tissue reduction along the network architecture may occur over time [16]; this putative ‘space-time interaction’ along the connectome structure can produce GMV variability despite a common mechanistic process operating across subjects. This will result in high variability of structural changes in early stages, but lower variability as the illness progresses and spatial convergence occurs. The contribution of connectomic architecture to anatomical heterogeneity in schizophrenia remains unexplored to date.

Brain network can serve as conduits for the spread of pathology, allowing illness processes originating in one region to propagate and affect distributed systems [17, 18]. The network-based ‘spreading’ pattern is initially observed in neurodegenerative disorders [19] and may also play a role in neuropsychiatric disorders such as schizophrenia [20, 21]. Recent studies show that cross-sectional cortical alterations across the course of schizophrenia strongly adhere to the organization of brain network architecture [20]. Another study also reports that longitudinal GMV changes in schizophrenia are indeed constrained by brain network architecture, consistent with expectations for a network-based ‘spreading’ pattern [21]. These evidences are based on group-level GMV difference between patients and controls; whereas inter-individual differences among patients are crucial for mapping network-based ‘spreading’ pattern at the individual level.

The first aim of the study is to comprehensively evaluate the brain structural heterogeneity in cross-sectional brain MRI data of 1,792 individuals with schizophrenia and address three specific questions. First, we tested for a systematic relationship between overall (mean) changes in GMV and variability in schizophrenia. Second, we tested if the anatomical heterogeneity is more pronounced in later stages of schizophrenia. Third, we tested if a putative diffusion process with GMV deficits originating in one part of the brain and spreading elsewhere explain the anatomical heterogeneity of schizophrenia. To this end, we quantified patients’ individual deviations from the normative level from 1,532 healthy controls and employed a network diffusion model [22] to simulate specific spatial patterning of GMV alteration. Additionally, we explored the association of brain variability in schizophrenia with neurotransmitter receptor expression distribution and transcriptomic gene expression data in the human brain. This allowed us to identify the processes contributing to brain structural alteration at an individual level, and unveiled the diversity of brain phenotypes with differential pathophysiological ‘sources’.

## 2. Methods

### 2.1 Sample

#### Discovery sample

The discovery sample included cross-sectional magnetic resonance imaging (MRI) T1-weighted scans from a total of 1,799 individuals diagnosed with schizophrenia (762 females, mean age=29.9±11.9 years) and 1,532 healthy controls (702 females, mean age=31.3±11.8 years). Details of demographics, clinical characteristics, and inclusion/exclusion criteria for each cohort are described in the **Supplementary Table 1-2**. All data obtained approval from their respective local institutional review boards or ethics committees, and written informed consent was obtained from all participants. This study was conducted under the approval of the Medical Research Ethics Committee of Fudan University.

#### External validation data

An external validation data included MRI-derived cortical thickness/subcortical volume statistical data in schizophrenia. Summary statistics (i.e., effect sizes for case-control differences) were obtained from ENIGMA data including over 4,000 scanned individuals with schizophrenia against almost 5,000 healthy controls in published studies [23, 24].

### 2.2 MRI-derived gray matter volume measurements

Brain images were processed by using FreeSurfer (http://surfer.nmr.mgh.harvard.edu/). Regional gray matter volume (GMV) was quantified by each of 68 cortical regions in the Desikan-Killian atlas [25], along with 14 subcortical regions including bilaterally thalamus, caudate, putamen, pallidum, hippocampus, amygdala and accumbens. Regional GMV measure was adjusted by regressing out the factors of no interest, such as sex, age, square of age, total intracranial volume (TIV) and site [5]. Sample outliers were removed if any of regional volume > 5 standard deviations away from the group-level average. A total of 3,315 subjects (1,792 patients) were included after quality control.

### 2.3 Variability measures in regional gray matter volume

To explore whether individuals with schizophrenia demonstrate higher variability of regional GMV, we computed the variability by comparing the relative variability of patient to control measures, using the log variability ratio (lnVR) as an index [1]. We used a relative mean-scaled variability (lnCVR), which accounts for differences in mean GMV [1]. The lnVR and lnCVR are given by the equation:

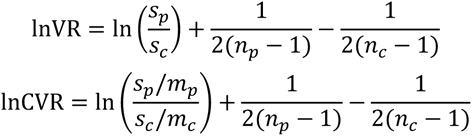

Where *s_p_* and *s_c_* are the sample standard deviations, *m_p_* and *m_c_* are the sample means, and *n_p_* and *n_c_* are the sample sizes for patient and control groups, respectively.

### 2.4 Individual deviation relative to healthy controls

We quantified GMV difference in each patient compared to the healthy control group by using *z score* [5]. Specifically, for each patient *i* and a given region *j*, the regional GMV measures were further transformed to *z scores* by the following:

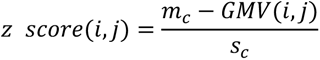

Where *m_c_* is the sample mean of control group, *s_c_* is the standard deviation of control group, and *GMV(i, j)* is the covariates-adjusted GMV of region *j* of patient *i*.

In this way, patient-specific GMV changes were estimated as *z scores*, which quantify the degree of deviations relative to an average of healthy controls. A higher *z score* for each patient signifies a greater deviation degree from the normal level, indicating a smaller GMV in this context. We also calculated the percentage of patients whose deviation was in the direction of reduction (PD−) or increase (PD+) in regional GMV compared to the normative levels (beyond the threshold of *z score*>1 or *z score*<-1).

### 2.5 Statistical analysis

#### 2.5.1 Patient-control comparison

Two sample *t* test was performed to compare the mean difference in covariates-adjusted regional GMV between patients and controls. Permutation test was used to compare the variability difference in regional lnCVR between patients and controls (**Supplementary Materials**). FDR correction was conducted for a total of 82 regional-level multiple comparisons.

#### 2.5.2 Spatial correlation

Spearman correlation tests were conducted to investigate associations (1) between two case-control mean difference (i.e., effect sizes) maps in discovery sample and validation sample; (2) between variability map (i.e., lnCVR) and effect size map; (3) between lnCVR and individual deviation to smaller GMV (i.e., PD−); and (4) between lnCVR and individual deviation to larger GMV (i.e., PD+). Spatial permutation test (i.e., *spin* test) was further employed for spatial autocorrelation adjustment (**Supplementary Materials**). FDR correction was utilized for multiple comparisons.

### 2.6 Transcriptomic analysis

We investigated the association between transcriptome and regional variability and individual deviation in schizophrenia by using Allen Human Brain Atlas dataset (AHBA) (http://human.brain-map.org). Details of transcriptomic analysis are described in **Supplementary Materials.** Briefly, partial least squares (PLS) correlation analysis [26] was used to examine the association of gene expression levels for 12,668 genes expressed in human brain with regional variability and individual deviation in schizophrenia. The associated genes with an FDR corrected *p*<0.05 was extracted for the gene enrichment analysis using Metascape [27].

### 2.7 Neurotransmitter receptors and transporters analysis

We investigated the relationships between regional variability (and individual deviation) in schizophrenia and neurotransmitter systems (Details are provided in **Supplementary Materials**). We used whole-brain positron emission tomography (PET) images across a cohort over 1200 healthy individuals [28]. Using these data, we examine the association of the lnCVR map (PD− and PD+ maps) in schizophrenia with neurotransmitter maps including dopamine (D_1_, D_2_, and DAT), norepinephrine (NAT), serotonin (5-HT_1a_, 5-HT_1b_, 5-HT_2a_, 5-HT_4_, 5-HT_6_, and 5-HTT), acetylcholine, glutamate (mGluR_5_ and NMDAR), gamma-aminobutyric acid (GABA_a_), histamine, cannabinoid, and opioid.

### 2.8 Network diffusion model

We employed a classical algorithm known as the NDM [22] to simulate the specific spatial patterning of GMV alterations through a putative diffusion process within a brain network. NDM has been applied in schizophrenia, revealing specific spatial patterning of GMV changes across different stages of psychosis [21]. Details of NDM are provided in **Supplementary Materials** and briefly described here. In NDM, pathological progression is assumed to be a diffusive propagation between connected brain regions, as described by the equation:

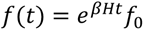

Where *t* is the model diffusion time, *f(t)* is a vector describing the pattern of diffusion in each region at time *t*, *β* is a diffusivity constant, and *H* is the Laplacian of the brain network, and *f_0_* is the initial distribution of pathology at *t*=0.

We used a seed-searching approach to detect the optimal seed for modeling. Specifically, we repeatedly initialized the model by using each of the 82 regions as the starting seed. We further computed the Spearman r values to determine whether such a diffusion process from specific seed region matched GMV changes from empirically observed data. An acceptable performance by NDM was defined as corrected *p_spin_*<0.001. In this way, it was able to determine the optimal connectome type and optimal seed region for NDM simulation of both group-level and individual-level GMV changes.

## 3. Results

### 3.1 Mean difference in regional gray matter measures

We found significant group differences of mean GMV in 73 brain regions (*p*<0.05, FDR corrected) (**Supplementary Table S3**). Most regional mean volumes were significantly lower in patients compared with healthy controls. Only bilateral pallidum volumes were significantly larger in patients. The regional effect sizes of group mean difference are mapped to a brain template (**Figure 1a**); the largest effect size is located at bilateral hippocampus (**Supplementary Table S3**). Consistent with another independent sample over 4000 individuals with schizophrenia, the effect size brain map showed a high spatial correlation (*r*=0.678, *p=*2.7×10^−12^) with the brain map of case-control difference collected from ENIGMA data (**Supplementary Figure 1**).

**Figure 1.**
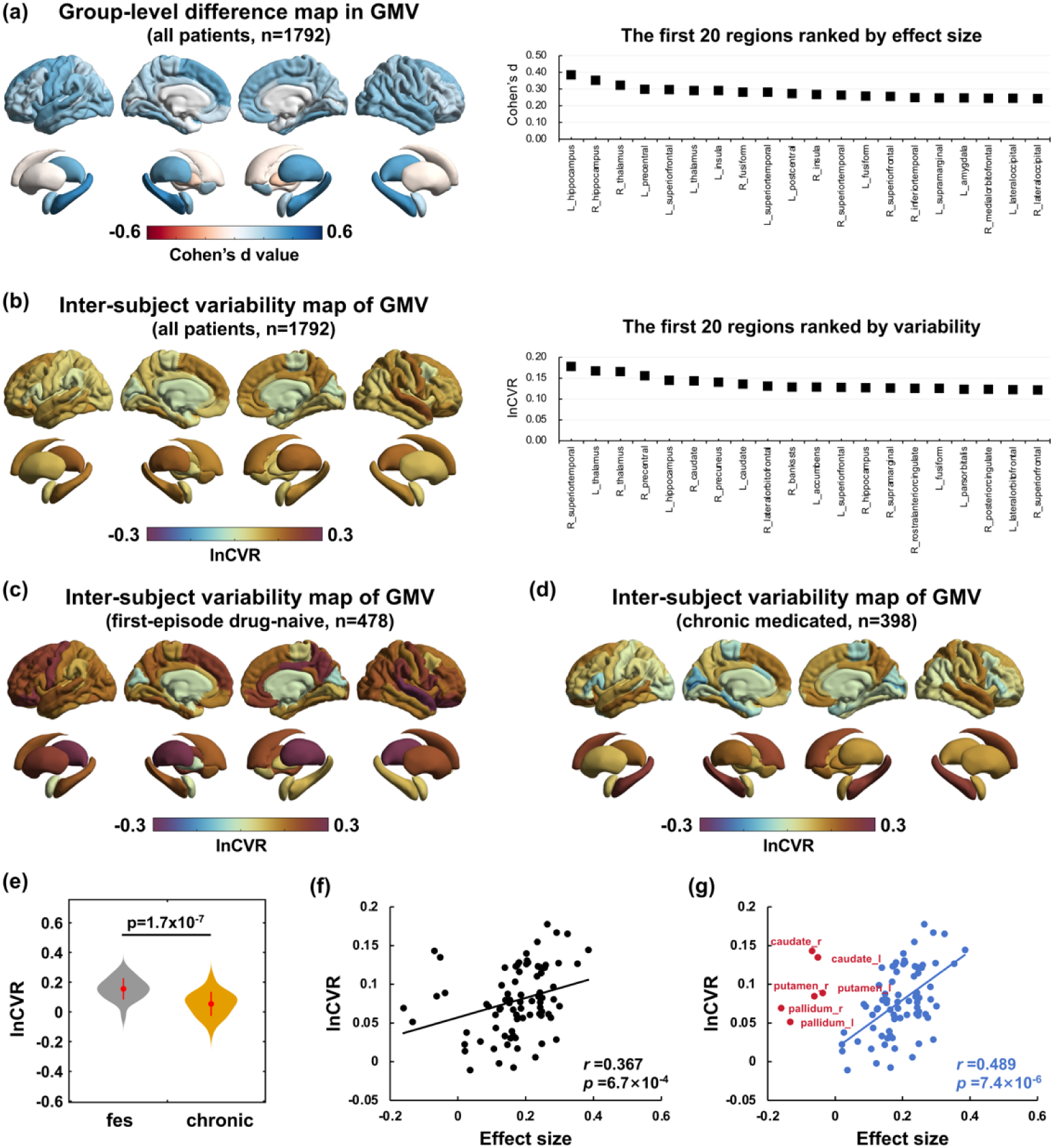
Group difference of mean volume and variability in regional gray matter (GM) measures between patients with schizophrenia and healthy controls. **(a)** Regional effect sizes of group difference in mean volume are mapped to a brain template. The right panel shows the first 20 brain regions ranked according to their effect sizes. **(b)** Regional variability values are mapped to a brain template. The variability is evaluated by comparing the relative variability of patients to controls, using a mean-scaled log variability ratio (lnCVR). The right panel shows the first 20 brain regions ranked according to their lnCVR values. **(c)** Spatial association between the lnCVR map and effect size map in case-control difference across whole brain regions (*r*=0.367, *p=*6.7×10^−4^). **(d)** The lnCVR map is significantly spatially correlated with effect size map across smaller volume regions (colored with blue) in patients relative to healthy controls (*r*=0.489, *p=*7.4×10^−6^).

### 3.2 Variability difference in regional gray matter measures

We calculated each regional lnCVR in the whole sample and two independent subsamples at early or late illness stages (first-episode drug-naïve subsample [n=478, age=23.1±7.6 years, 239 females, illness duration=0.548±0.459 years] and chronic medicated subsample [n=398, age=37.9±12.1 years, 139 females, illness duration=14.0±10.4 years]). **Figure 1b-d** show regional lnCVR values across all brain regions by mapping them to a brain template. Compared with healthy controls, significant greater lnCVR (*p*<0.05, FDR corrected) was found in 50 regions in the whole patient group, at a much greater frequency in the first-episode group (73 regions), compared to the chronic group (28 regions; Chi-square test, *p=5.0*×10^−13^). The greater lnCVR in first-episode than chronic group was also replicated in females (*p=2.7*×10^−21^) or males (*p=9.5*×10^−8^) (**Supplementary Materials**). The areas with largest variability were mainly located at the frontotemporal and thalamus for first-episode patients, or the hippocampus and caudate for chronic patients (**Figure 1b-d**). Significantly lower variability than controls was not found in any regions in the whole patient group or in the first-episode group, but in bilateral pericalcarine, left cuneus and left parahippocampal in the chronic subsample (*p*<0.05, FDR corrected). In addition, the average lnCVR across all regions was higher in the first-episode subsample than chronic subsample in a head-to-head comparison (*t*=10.8, *p=*1.7×10^−7^) (**Figure 1e**). **Supplementary Table S4** provides statistical results of regional-wise case-control comparison in each group. We also derived a score by ranking ratio between the effect size in group-level mean difference and variability for each region to identify regions that are ‘invariant’ in gray matter reduction (**Supplementary Materials**).

### 3.3 Individual deviation from the normal level

We measured the individual deviation, by quantifying the proportion of patients whose regional GMV exhibited an ‘abnormal’ deviation to reduction (PD−) or increase (PD+) relative to the average of healthy people (**Figure 2a and Supplementary Table S5**). The highest proportions of patients had PD− at bilateral thalamus (27.0% patients in the right and 26.1% in the left thalamus (**Figure 2b**)) and PD+ at basal ganglia sub-regions. Specifically, 19.3%, 18.8% and 18.0% patients showed individual-level volume increase in the right pallidum, right and left caudate, respectively (**Figure 2c**). We also computed PD− and PD+ in first-episode drug-naïve subsample and chronic subsample (**Supplementary Table S5**).

**Figure 2.**
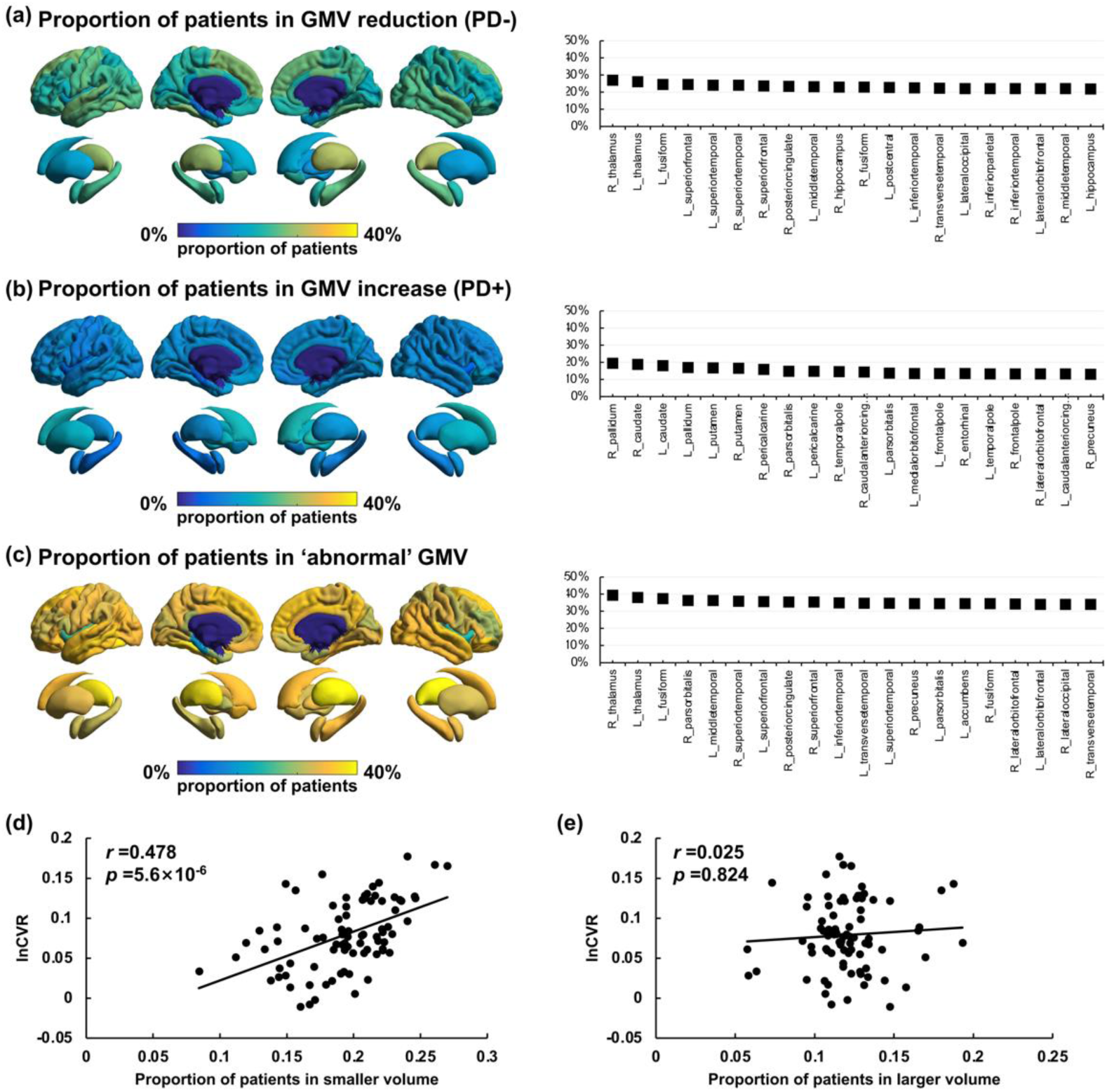
Individual-level deviation from the normal level in regional gray matter volume (GMV) changes. Regional GMV difference in each patient compared to the healthy control group is quantified by using z score. The left panels show the proportion of patients for each brain region with a deviation (more than one standard deviation from the average of normal population) of **(a)** smaller volume, **(b)** larger volume, and **(c)** both ‘abnormal’ volume. The right panels show the first 20 brain regions ranked according to their proportion values. **(d)** The variability (lnCVR) map is significantly spatially correlated with the brain map in proportion of patients in smaller volume (*r*=0.478, *p=*5.6×10^−6^), **(e)** but not with the brain map in proportion of patients in larger volume (*r*=0.025, *p=*0.824).

### 3.4 Spatial correlation between regional variability and volume reduction

We found that the lnCVR map was significantly spatially correlated with effect size map in case-control difference (*r*=0.367, *p=*6.7×10^−4^) (**Figure 1f**). The first 10 regions with highest lnCVR (except bilateral caudate regions) showed obvious volume reduction in patients (**Supplementary Table S4**). Furthermore, it showed a stronger spatial correlation (*r*=0.489, *p=*7.4×10^−6^), when correlation test was limited on these regions with smaller volume (i.e., Cohen’s d>0) in patients relative to controls (**Figure 1g**). In addition, the lnCVR was significantly spatially correlated with the PD− map (*r*=0.478, *p=*5.6×10^−6^) (**Figure 2d**), but not with the PD+ map (*r*=0.025, *p=*0.824) (**Figure 2e**). Details of spatial correlation results are provided in **Supplementary Table S6**.

### 3.5 Molecular mechanisms of regional variability in schizophrenia

We further conducted transcriptomic analysis to determine which genes were associated with regional lnCVR and PD− (PD+) in schizophrenia. We found that the spatial map of PLS1 (**Figure 3a**) was positively correlated with the lnCVR map (*r*=0.491, *p*=0.003; **Figure 3b**). We ranked the genes according to their weights to PLS1 (**Figure 3c**), resulting in a total of 829 significant genes (FDR *p*<0.05) for enrichment analysis, which revealed the top 20 biological processes (**Figure 3d**). In cell type signatures, these genes were enriched in the multiple neuron types, such as GABAergic, serotonergic and dopaminergic neurons (**Figure 3e**). Human disease-associated gene enrichment analysis showed that these genes are mainly enriched in the hypoplasia of corpus callosum (**Figure 3f**).

**Figure 3.**
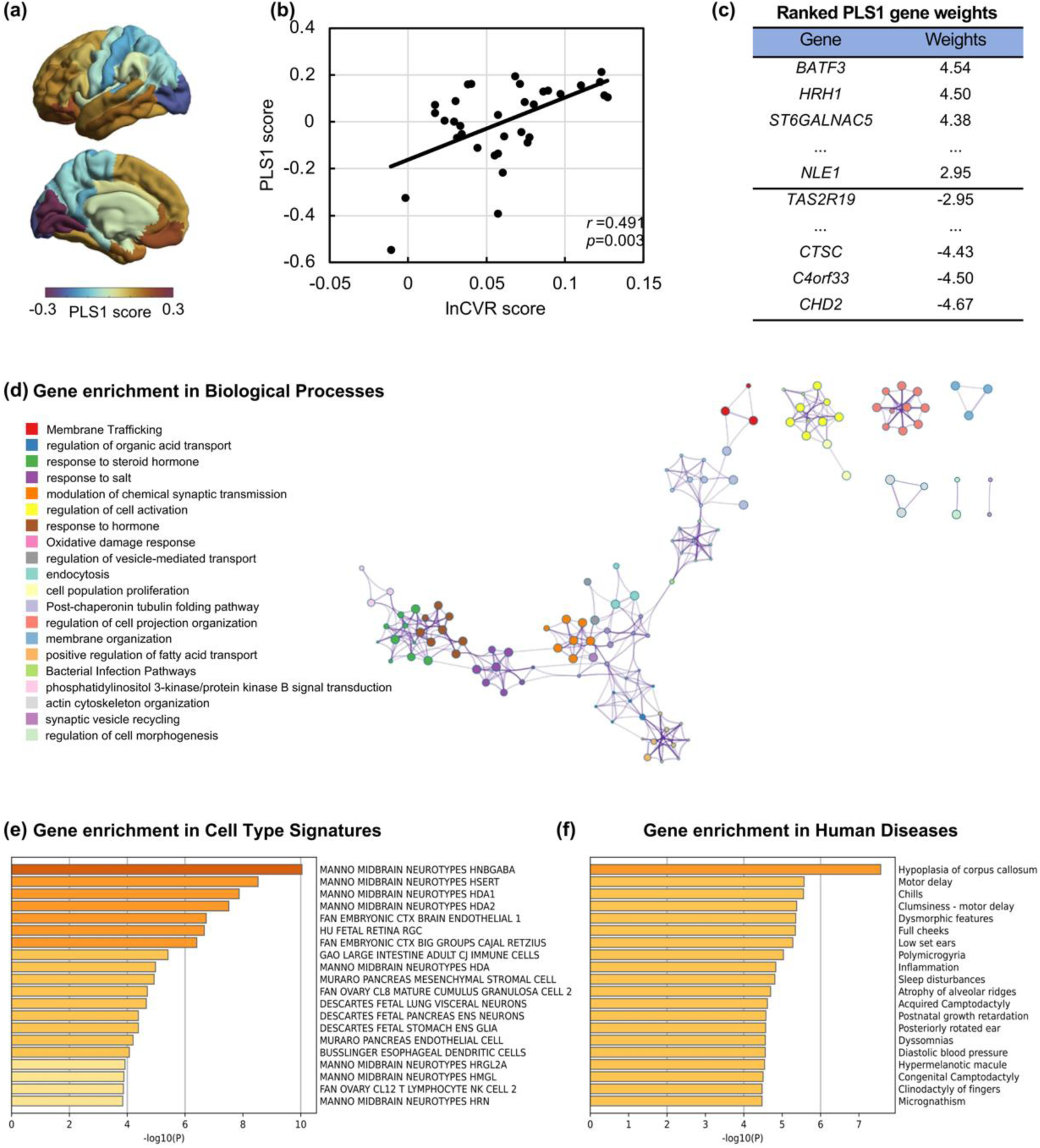
Partial least squares (PLS) correlation analysis between regional variability and human brain gene expression data. **(a)** Spatial map of the first component (PLS1) in the left hemisphere. **(b)** The spatial correlation between the PLS1 score and the variability in schizophrenia, measured by lnCVR score. **(c)** The ranked gene weights of PLS1 (FDR *p*<0.05). **(d)** Top 20 gene ontology (GO) biological processes by enrichment analysis. Circle color: different biological processes. Circle size: number of genes. **(e)** Gene enrichment analysis in Cell Type Signatures. **(f)** Gene enrichment analysis in human disease-associated database.

There was a spatial association (*r*=0.391, *p*=0.022) between the PD− map and gene PLS1 map (see **Supplementary Figure 2 and Supplementary Materials**). There was no significant spatial correlation between PD+ map and gene PLS1 map (*r*=0.216, *p*=0.219). In addition, we found significant correlations between brain variability map and neurotransmitter maps (**Supplementary Table 7**).

### 3.6 Network-based diffusion modeling at group-level and individual-level

We next employed the NDM to simulate the possible processes (i.e., spatial ‘spreading’ pattern) of GMV alteration across brain regions. We found that the group-level GMV difference in patients was successfully simulated by using the NDM in both discovery and validation samples (**Supplementary Figure 3 and Supplementary Table 8**). Temporal lobe regions (mainly the hippocampus) were consistently identified as the optimal source seed, across the three models, and across subsamples at different illness stages (first-episode, drug-naïve, and chronic) (**Supplementary Figure 3**). A full list is provided in **Supplementary Table 9** to show the model performances with each region being the starting point of NDM. This indicates that ‘spreading’ atrophy patterns starting at hippocampus, pars opercularis and thalamus are likely explanations for the variability seen in patients.

At the individual-level, we found that the NDM can achieve an accepted performance (*p_spin_*<0.001) in the consistency between estimated values and observed values for 72.1% patients (SZ_NDM_, n=1,292) (**Supplementary Table 10a**). For these SZ_NDM_ patients, we further classified them into several subgroups based on the heterogeneity in optimal seed locations (**Supplementary Table 10b**). **Figure 4** shows specific spatial patterns of GMV change in each spatial phenotype, indicating that ‘temporal cortex’ and ‘frontal cortex’ phenotypes account for the highest proportion of patients, reaching 24.2% and 13.6% respectively. **Supplementary Table 11** shows regional comparison of GMV between patients and controls for each spatial phenotype. A detailed description is provided in **Supplementary Materials.** Together, we show the existence of neuroanatomically distinct phenotypes, which are characterized with differential spatial patterns of gray matter alterations in schizophrenia.

**Figure 4.**
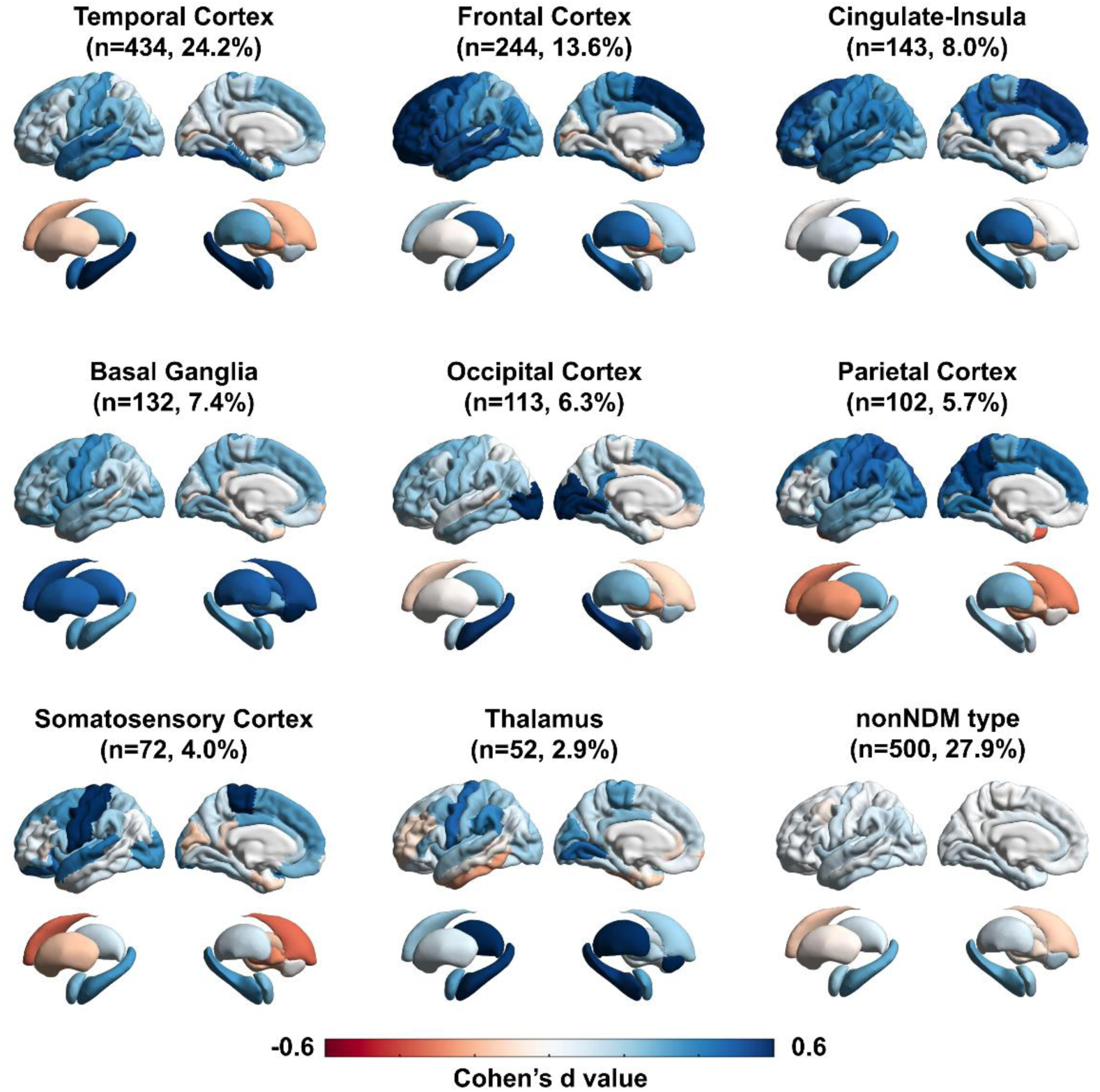
Distinct neuroanatomical characteristics in network diffusion model (NDM)-based ‘spatial’ phenotypes of schizophrenia population. Patients are assigned to subgroups (termed as ‘spatial’ phenotypes) based on the heterogeneity in their optimal NDM seed locations. Spatial patterns of specific gray matter change are shown by mapping case-control effect sizes to a brain template in each ‘spatial’ phenotype, which are labelled as ‘temporal cortex’, ‘frontal cortex’, ‘occipital cortex’, ‘parietal cortex’, ‘somatosensory cortex’, ‘cingulate-insula’, ‘basal ganglia’ or ‘thalamus’ phenotypes according to an anatomical definition. In addition to the nine ‘spatial’ phenotypes, patients who are not estimated by NDM are assigned to another phenotype (‘nonNDM’). The number and proportion of patients in each phenotype are also described. Color bar represents the case-control effect size measure by Cohen’s d value. A larger Cohen’s d value indicates severer reduction in gray matter volume.

## 4. Discussion

Our work assessed the inter-subject variability of regional GMV in schizophrenia. We found support for a space-time interaction along a shared pathophysiological continuum (network-based trans-neuronal diffusion), as a possible explanatory model for inter-subject variability. This allowed us to delineate the potential genetic and neurotransmitter basis for brain structural variability in schizophrenia without invoking the need for mechanistic subgroups. These findings contribute to the understanding that inter-individual variability in schizophrenia may arise from a common cohesive process that varies in its state (across time) and space (across brain regions).

One of our main findings is that inter-individual differences of brain structure are vast in people diagnosed as schizophrenia. An earlier study has reported that schizophrenia is associated with greater variability (i.e., heterogeneity) of temporal cortex, thalamus and putamen [1], but this was limited to meta-analytical data. In addition to these regions [1], our individual level data found significant greater variability in more brain areas, including frontotemporal structures, hippocampus, basal ganglia and other sub-cortical regions. Interestingly, for the first time, we show the relationship between greater variability and illness stages, pointing out the exaggerated or hyper-variability in the first-episode drug-naïve subsample, which is relatively less pronounced (despite significantly higher than controls) in patients with longer term illness. Our observations argue against the expectations [7] that inter-individual variability may increase in established cases as patients may vary across illness stages and severity, due to differences in treatment status, varied progression and ageing.

The relatively large hyper-variability at the disease onset may imply either the numerosity of factors contributing to gray matter changes or the multiplicity of response states emerging from the converging effects of the various mechanistic factors in schizophrenia [29]. The latter explanation is supported by the observation that hyper-variability is more likely in those regions that show a shared mean reduction in patients. Furthermore, over a longer term, a reduction of hyper-variability with the emergence of hypo-variability in some regions occurs, likely a result of the convergence of the diverse etiogenic factors on a common path, thus reducing heterogeneity. The regional distribution of lower variability in schizophrenia (i.e., homogeneity) has been inconsistent in prior studies [1, 7, 30] (see more discussion in **Supplementary Materials**).

Another finding shows that the inter-patient variability is related to several neurobiological processes closely linked to the etiology of schizophrenia during neurodevelopmental processes. Our data show that: (1) the genes whose expression is spatially associated with lnCVR in schizophrenia were mainly enriched for biological processes related to synaptic functions. Synaptic dysfunction is a long-standing hypothesis [31–33], holding the aberrant synaptic pruning during development as the etiology of schizophrenia. (2) Genes associated with lnCVR were also enriched in the midbrain GABAergic, serotonergic and dopaminergic neuron types, which have been demonstrated be involved in the etiology of schizophrenia [34]. This is also consistent with prior research elucidating multi-cellular correlates of cortical thinning in schizophrenia [35]. (3) lnCVR-related gene enrichment in human diseases points out hypoplasia of corpus callosum. The etiopathology of hypoplastic corpus callosum [36, 37] is related to atypical brain connection, which has been generalized as ‘dys-connection syndrome’ in discussing the pathogenesis of schizophrenia [38, 39]. (4) Final, the high variability regions in first-episode schizophrenia were primarily located within frontotemporal cortex, a region that is said to be affected as a result of aberrant neurodevelopmental processes relevant to the early stage of schizophrenia [5, 40]. Together, our study supports diversity in the etiology of schizophrenia, and suggests that multiple biological processes involved in neurodevelopment may ultimately manifest as individual differences in brain mesoscale structure in schizophrenia.

Our modeling algorithm shows the potential to uncover the heterogeneity in pathophysiological processes of GMV change in schizophrenia. First, the group-level modeling identified the temporal lobe structure (mainly the hippocampus) as an optimal putative source of GMV change in schizophrenia. It was also replicated using independent ENIGMA data, and consistent with a prior NDM modeling in psychosis [21]. The hippocampus has been consistently implicated in the pathogenesis of schizophrenia [41, 42] (also see [43]), highlighting hippocampal dysfunction during the early stages of the disease [44, 45]. Recent research suggests that dysregulation of glutamate neurotransmission, which initially occurs in the hippocampal CA1, leads to atrophy in other medial temporal areas and their connected regions [41, 46]. Furthermore, Lower volume in the CA1 is predictive of the transition to psychosis within a two-year timeframe [40]. Positron emission tomography (PET) imaging has also revealed a reduction in synaptic vesicle proteins in the hippocampus in schizophrenia [47]. In essence, the highly consistence of modeling results from two independent dataset demonstrates the robustness of NDM algorithm and reproducibility of hippocampus role as the ‘source’ of group-level GMV change in schizophrenia.

In contrast, the individual-level modeling identifies the heterogeneity of pathophysiological processes, which can be characterized into several differential GMV change patterns (termed as ‘spatial’ phenotypes). This finding aligns with a recent study, which indicates distinct ‘sources’ of GMV losses related to disease stage, longitudinal progression, or antipsychotic medication [21]. The ‘sources’ of illness-related GMV loss were found in the hippocampus and the prefrontal cortex; whereas the ‘sources’ of antipsychotic-related loss were identified in somatosensory, motor, and cingulate regions [21]. Although the exact cause of difference among ‘spatial’ phenotypes is unclear, the result provides direct brain imaging evidence supporting the existence of brain phenotypic heterogeneity in pathophysiological processes of schizophrenia. These phenotypes could shed light on primary or common biological characteristics in the pathological mechanisms of schizophrenia, which may help delineate potential subtypes of this disorder. We also provide an approach to detect homogeneous people with shared focal ‘lesion’ for drug development and intervention target.

The large sample size used for examining variability is a strength of this work; the inclusion of samples from various sites improves generalizability of findings across diverse cohorts, scanners, or locations. Despite this, our method has some limitations. While we employed harmonization procedures to reduce potential bias, it is essential for future multi-site collaborations use a standard protocol. Second, we related the inter-subject variability map in schizophrenia with the gene expression brain map sourced from healthy individuals. The observed spatial association still needs to be verified on patient level genetic data. Finally, the NDM modeling was constructed based on cross-sectional data. Longitudinal studies are better suited for making inferences on ‘propagation’ or spreading of changes with the temporal progression of pathophysiological processes.

In conclusion, this study offers novel insights into the diversity of neuroanatomical alterations in schizophrenia, emphasizing that brain-based heterogeneity is not a static feature of schizophrenia; it is more pronounced at the onset of the disorder but reduced over the long term. This heterogeneity is associated with various molecular and neurobiological processes implicated in the neurodevelopmental etiology of schizophrenia. Thus, the molecular etiological diversity, influencing neuroanatomical sites of origin of gray matter reduction in each individual with schizophrenia, may ultimately manifest as variability in brain’s mesoscale structure among patients. By presenting an approach to understand heterogeneity through individually distinct pathophysiological ‘sources’ of changes, our work also raises the question of what dynamic processes contribute to the reducing heterogeneity over time in schizophrenia. Answering this question will be a key test to the neurobiological validity of the concept of schizophrenia.

## Author contributions

Concept and design: Yuchao Jiang, Jianfeng Feng.

Acquisition of data: Jie Zhang, Enpeng Zhou, Xin Yu, Shih-Jen Tsai, Ching-Po Lin, Jingliang Cheng, Yingying Tang, Jijun Wang, Cheng Luo, Dezhong Yao, Long-Biao Cui.

Analysis, or interpretation of data: Yuchao Jiang, Xiao Chang.

Original drafting of the manuscript: Yuchao Jiang, Lena Palaniyappan.

Critical review of the manuscript for important intellectual content: Lena Palaniyappan, Xiao Chang, Wei Cheng.

Statistical analysis: Yuchao Jiang, Xiao Chang. Supervision: Jianfeng Feng.

## Data Availability

All data produced in the present study are available upon reasonable request to the authors.

## Acknowledgements

This work was supported by the grant from Science and Technology Innovation 2030-Brain Science and Brain-Inspired Intelligence Project (No. 2022ZD0212800 to Y.J.). This work was supported by National Natural Science Foundation of China (No. 82202242 to Y.J.; No. 82071997 to W.C.). This work was supported by the projects from China Postdoctoral Science Foundation (No. BX2021078 and 2021M700852 to Y.J.). This work was supported by the Shanghai Rising-Star Program (No. 21QA1408700 to W.C.) and the Shanghai Sailing Program (22YF1402800 to Y.J.) from Shanghai Science and Technology Committee. This work was supported by National Key R&D Program of China (No. 2019YFA0709502 to J.F.), the grant from Shanghai Municipal Science and Technology Major Project (No. 2018SHZDZX01 to J.F.), ZJ Lab, and Shanghai Center for Brain Science and Brain-Inspired Technology, and the grant from the 111 Project (No. B18015 to J.F.). This work was supported by grants from the National Key R&D Program of China (No. 2022ZD0208500 to D.Y.) and the CAMS Innovation Fund for Medical Sciences (No. 2019-I2M-5-039 to C.L.). LP acknowledges research support from the Canada First Research Excellence Fund, awarded to the Healthy Brains, Healthy Lives initiative at McGill University (through New Investigator Supplement to LP); Monique H. Bourgeois Chair in Developmental Disorders and Graham Boeckh Foundation (Douglas Research Centre, McGill University) and a salary award from the Fonds de recherche du Quebec-Santé (FRQS).

## Data availability

Data of COBRE, NMorphCH, FBIRN and NUSDAST were obtained from the SchizConnect, a publicly available website (www.schizconnect.org). The COBRE dataset was download from the Center for Biomedical Research Excellence in Brain Function and Mental Illness (COBRE) (http://coins.mrn.org/). The NMorphCH dataset was download from https://nunda.northwestern.edu/nunda/data/projects/NMorphCH. The FBIRN dataset was download from https://www.nitrc.org/projects/fbirn/. The NUSDAST dataset was download from the Northwestern University Schizophrenia Data and Software Tool. The DS000115 dataset was download from OpenfMRI database (https://www.openfmri.org/). ENIGMA summary statistics of thinner cortical thickness map were obtained from ENIGMA toolbox (https://github.com/MICA-MNI/ENIGMA) (version 2.0.0, July, 2022). All data needed to evaluate the conclusions in the paper are present in the paper and/or the Supplementary Materials.

## Code availability

Brain images were processed by using FreeSurfer (http://surfer.nmr.mgh.harvard.edu/). Gene enrichment analysis was performed using Metascape (https://metascape.org). The visualization of brain mapping images was conducted using ENIGMA Toolbox (https://enigma-toolbox.readthedocs.io/en/latest/index.html). The code for spatial permutation test is available at (https://github.com/frantisekvasa/rotate_parcellation).

