## Supplementary Materials for "Brain heterogeneity in 1,792 individuals with schizophrenia: effects of illness stage, sites of origin and pathophysiology"

Yuchao Jiang, et al

**This supplementary material includes as follows:**


#### Part 1: Methods

Method S1. Null model and permutation test.

Method S2. Transcriptomic analysis.

Method S3. Neurotransmitter receptors and transporters analysis.

Method S4. Network diffusion model.

Method S5. Brain connectome estimation.

#### Part 2: Results

Result S1. Regional variability in females and males.

Result S2. Identifying regions that are 'invariant' in gray matter reduction.

Result S3. Association between PD- map and human brain gene map.

Result S4. Association between regional variability in schizophrenia and neurotransmitter systems.

Result S5. Network-based diffusion modeling at group-level and individual-level.

#### Part 3: Discussion

Discussion S1. Lower variability in schizophrenia is inconsistent in prior studies.

#### Part 4: Supplementary Tables

Table 1. Sample demographics by participating cohort.

Table 2. Dataset-specific information.

Table 3. Mean difference in regional gray matter volume measures.

Table 4. Variability difference in regional gray matter volume measures.

Table 5. Proportion of patients with greater deviation in regional gray matter volume measures.

Table 6. Spatial correlation results.

Table 7. Correlation between brain variability map in schizophrenia and neurotransmitter map.

Table 8. Network diffusion model performances on estimating cross-sectional gray matter differences in schizophrenia.

Table 9. Model performance with each brain region as the seed of network diffusion model.

Table 10. Network diffusion model (NDM)-based ‘spatial’ phenotypes in schizophrenia population.

Table 11. Regional comparisons of gray matter volume between each spatial phenotype in schizophrenia and healthy controls.

**Part 5: Supplementary Figures**

Supplementary Fig.1. High consistency between the case-control difference (i.e., effect size) brain map that sourced from (a) discovery sample and (b) validation sample.

Supplementary Fig.2. Partial least squares (PLS) correlation analysis between individual deviation and human brain gene expression data.

Supplementary Fig.3. Performance of network diffusion model (NDM) on estimating cross-sectional GMV differences in group-level patients with schizophrenia.

### Method S1. Null model and permutation test

#### 1. Permutation statistical test

To examine whether the variability of regional volume (lnCVR) was significantly different between the patient group and healthy control group, we used a permutation-based test. First, we computed the true lnCVR based on the true labels of patient or control. Second, a permutation step was conducted by randomly swapping labels of observation, and generated a possible lnCVR. The permutation process was repeated 1,000 times, and thus generated a distribution of the possible values under the null hypothesis that observations are exchangeable. The distributions of possible values among 1,000 permutations in each brain region exhibit normal distributions. According to the location of true value in the distribution of the possible values, the P-value (two-sided) was estimated to be how probable such observations would be if the null hypothesis is true.

#### 2. Spatial permutation test

We utilize the spin test, specifically the spatial permutation test, to address spatial autocorrelation when assessing the correspondence between the brain epicenter map and other maps [1, 2]. This approach is increasingly acknowledged as a leading method for examining spatial concordance in human brain mapping and has been extensively applied in numerous neuroimaging investigations [3-5].

A recent study has conducted a comprehensive analysis of ten null models for evaluating the correspondence of brain maps in neuroimaging research [6]. The findings reveal that naive null models, which neglect to preserve spatial autocorrelation, consistently lead to higher false positive rates and unrealistically liberal statistical estimates [6]. Additionally, the study demonstrates significantly enhanced performance and decreased error rates in null models that account for spatial autocorrelation compared to spatially-naive models [6]. In summary, based on these comprehensive comparisons, there is a strong recommendation for the adoption of spatial permutation null models [6].

The spin test employed here is a spatial permutation method based on angular permutations of spherical projections at the cortical surface. Critically, the spin test preserves the spatial covariance structure of the data and as such is far more conservative than randomly shuffling locations, which destroys the spatial covariance structure and produces an unrealistically unconservative null distribution. A previous study indicated that null models tend to converge and yield stable statistical estimates after generating approximately 100 to 500 nulls. Here, we generated 10,000 random spatial rotations (i.e., spins) of the brain regions to generate a null distribution. Then, the $p_{spin}$ values were obtained according to the location of true value within the null models.

### Method S2. Transcriptomic analysis

**Transcriptomic data**

The Allen Human Brain Atlas dataset (AHBA) (http://human.brain-map.org) provides gene expression data of brain tissue samples obtained from six postmortem human brains, which allow to investigate the association between transcriptome and regional variability and individual deviation in schizophrenia. After preprocessing AHBA data using the *abagen* [7], regional microarray expression data were extracted using the same regions with above GMV measurements. To get stably expression, genes exhibiting a low level of similarity across donors ($r$<0.2) were removed, resulting in a final set of 12,668 genes. Since only two donors have available data in the right hemisphere, we used the left hemisphere gene expression data for the following analyses.

**Partial least squares correlation analysis**

Partial least squares (PLS) correlation analysis [8] was used to examine the association of gene expression levels for all 12,668 genes with regional variability and individual deviation in schizophrenia. First, regional-wise gene expression data were used as predictor variables, and regional variability (lnCVR) or individual deviation (PD- or PD+) values were used as the response variables in PLS correlation. The first component of PLS (PLS1) indicated a strong spatial correlation with response variables. To correct spatial autocorrelation, spatial permutation test (*spin* test) was employed, as detailed in the **Supplementary Materials**. A boostrapping method was conducted to estimate the variance in the weight of each gene in the PLS analysis. The ratio of the weight of each gene to its bootstrap standard error (iterations=1000) was utilized to compute a Z score, allowing for the ranking of genes according to their contribution to PLS1 [9]. The list of genes with an FDR corrected $p$<0.05 was extracted for subsequent enrichment analysis.

**Gene enrichment analysis**

Gene enrichment analysis was performed using Metascape [10]. The genes list generated from PLS analysis were input into the Metascape analysis. Pathway and process enrichment analysis was carried out with the ontology sources: KEGG Pathway, GO Biological Processes, Reactome Gene Sets, Canonical Pathways, CORUM, WikiPathways, and PANTHER Pathway. P values were calculated based on the cumulative hypergeometric distribution. The Benjamini-Hochberg procedure was used to calculate q values account for multiple tests [11].

#### Method S3. Neurotransmitter receptors and transporters analysis

We investigated the relationships between regional variability (and individual deviations) in schizophrenia and neurotransmitter systems across all brain regions. To this end, we used whole-brain positron emission tomography (PET) images across a cohort over 1200 healthy individuals, sourced from multiple studies (accessible at https://github.com/netneurolab/hansen_receptors). Each of previous studies provided the mean tracer map across subjects, with details including the associated receptor/transporter, tracer used, number of healthy participants, age, and reference with full methodological details, as summarized in a previous study [5]. Using these data, we obtained a comprehensive profile of neurotransmitter receptors and transporter densities in brain, including 19 distinct neurotransmitter receptors and transporters across nine different neurotransmitter systems. Specifically, PET images were initially registered to a normalization template (MNI-ICBM 152 nonlinear 2009 template) and subsequently parcellated into 68 cortical and 16 subcortical regions, using the same atlases with the gray matter volume measurement. These images represent estimates proportional to receptor densities and are therefore referred to simply as 'density' here. Subsequently, the density values of each neurotransmitter receptor/transporter were transformed into z-scores across all regions. This process yielded a 84x19 matrix representing relative neurotransmitter receptor/transporter densities across 19 different neurotransmitter systems, including dopamine ($D_1$, $D_2$, and DAT), norepinephrine (NAT), serotonin ($5\text{-}HT_{1a}$, $5\text{-}HT_{1b}$, $5\text{-}HT_{2a}$, $5\text{-}HT_4$, $5\text{-}HT_6$, and 5-HTT), acetylcholine, glutamate ($mGluR_5$ and NMDAR), gamma-aminobutyric acid (GABAa), histamine, cannabinoid, and opioid. Next, Spearman correlation analysis was conducted to explore the spatial relationship between the regional variability map (and individual deviation maps) in schizophrenia and the neurotransmitter receptor/transporter map across these brain regions. FDR correction was used for multiple comparisons correction. Spin tests were utilized to correct for spatial autocorrelation.

#### Method S4. Network diffusion model

We employed a classical algorithm known as the NDM [12] to simulate the specific spatial patterning of GMV alterations through a putative diffusion process within a brain network. It enabled us to identify potential focal points of brain structural alteration. NDM has been applied in schizophrenia, revealing specific spatial patterning of GMV changes across different stages of psychosis [13].

##### 2.8.1 Brain connectome estimation

Brain connectivity data were derived from MICA-MICs dataset [14]. Normative brain connectome architectures were estimated from diffusion MRI, resting-state functional MRI, and T1-weighted MRI acquired in an independent sample of 50 healthy participants (23 women; mean age=29.54±5.62 years). Multi-modal MRI data were processed using the open-source pipeline, micapipe (https://micapipe.readthedocs.io). We obtained multi-modal connectomes including structural connectivity (SC), functional connectivity (FC) and morphological covariance connectivity (MC), generated from the diffusion scans, resting-state functional scans and T1-weighted scans separately. A detailed description of the methods to generate the connectomes is described in **Supplementary Materials**. All connectomes were generated from the 68 cortical regions in the Desikan-Killian atlas [15] and 18 subcortical regions, for consistency with GMV measurements. Group-averaged connectomes across participants were used as the reference brain connectivity for the following NDM.

##### 2.8.2 Network diffusion modeling

We used NDM to test whether GMV change spreads through the brain via a process of diffusion. In NDM, pathological progression is assumed to be a diffusive propagation between connected brain regions, as described by the equation:

$$f(t) = e^{\beta H t} f_0$$

Where $t$ is the model diffusion time, $f(t)$ is a vector describing the pattern of diffusion in each region at time $t$, $\beta$ is a diffusivity constant, and $H$ is the Laplacian of the brain network, and $f_0$ is the initial distribution of pathology at $t$=0.

We used a seed-searching approach to detect the optimal seed for modeling. Specifically, we repeatedly initialized the model by using each of the 82 regions as the starting seed. In this process, the initial state $f_0$ was set to 1 for the seed region (bilateral) and 0 for all other regions [12]. During each initialization, with a constant value of $\beta$=1, we estimated the diffusion $f(t)$ at all other regions from time $t$= 1 to 50 [12]. We further determined whether such a diffusion process from specific seed region matched GMV changes from empirically observed data. We calculated the Spearman r values between the spatial pattern of empirically observed GMV changes and the NDM estimated patterns. Spatial permutation test (i.e., *spin* test) was utilized to address spatial autocorrelation correction. An acceptable performance by NDM was defined as corrected $p_{spin}$<0.001. In this way, it was able to determine the optimal connectome type and optimal seed region for NDM simulation of both group-level and individual-level GMV changes.

#### Method S5. Brain connectome estimation

Brain connectivity data were derived from MICA-MICs dataset [14]. Normative brain connectome architectures were estimated from diffusion MRI, resting-state functional MRI, and T1-weighted MRI acquired in an independent sample of 50 healthy participants (23 women; mean age=29.54±5.62 years). Multi-modal MRI data were processed using the open-source pipeline, micapipe (https://micapipe.readthedocs.io). We obtained multi-modal connectomes including structural connectivity (SC), functional connectivity (FC) and morphological covariance connectivity (MC), generated from the diffusion scans, resting-state functional scans and T1-weighted scans separately.

##### 1. Structural connectivity (SC)

Structural connectomes were constructed using MRtrix from pre-processed diffusion MRI data [16]. Anatomically-constrained tractography was performed using tissue types segmented from each participant's pre-processed T1-weighted (T1w) images, which were registered to native DWI space [17]. Multi-shell and multi-tissue response functions were estimated, followed by constrained spherical deconvolution and intensity normalization [18]. A tractogram comprising 40 million streamlines (with a maximum tract length of 250 and a fractional anisotropy cutoff of 0.06) was generated. Spherical deconvolution informed filtering of tractograms (SIFT2) was applied to reconstruct whole brain streamlines weighted by cross-sectional multipliers [19]. The reconstructed streamlines were then mapped to each parcellation scheme (both cortical and subcortical), which were also aligned to DWI space. The connection weights between nodes were defined based on the weighted streamline count.

##### 2. Functional connectivity (SC)

Resting-state functional MRI time series, were firstly mapped to subject-specific surface models, and subsequently averaged within cortical parcels. The subcortical parcellation was also aligned to each subject's native functional MRI volume space and used to average time series within each node. Individual functional connectomes were generated by cross-correlating all nodal time series. Correlation coefficient values underwent Fisher's r-to-z transformations.

##### 3. Morphological covariance connectivity (MC)

Morphological covariance network was generated from T1-weighted images in the dataset. The FreeSurfer processing pipeline was used to calculate the cortical volume values for each region in the Desikan-Killiany atlas and subcortical volume. Linear regression was performed for each region to remove the effects of age, gender, and TIV; the residuals of this regression were used for a Pearson’s correlation analysis. Connectivity weights were defined as the z transformation of Pearson’s R value across subjects; Regions are considered connected if their regional volumes co-vary across subjects.

#### Result S1. Regional variability in females and males

We also calculated regional lnCVR in female patients and male patients separately. Differences of lnCVR between patients and healthy controls are provided in the below table 1.

In all people (females and males), significant greater lnCVR ($p$<0.05, FDR corrected), compared with healthy controls, was found in 50 regions in the whole patient group, at a much greater frequency in the first-episode group (73 regions), compared to the chronic group (28 regions; Chi-square test, $p=5.0\times10^{-13}$).

In females, we found that compared with healthy controls, significant greater lnCVR ($p$<0.05, FDR corrected) was found in 17 regions in the whole patient group, at a much greater frequency in the first-episode group (65 regions), compared to the chronic group (5 regions; Chi-square test, $p=2.7\times10^{-21}$).

In males, we found that compared with healthy controls, significant greater lnCVR ($p$<0.05, FDR corrected) was found in 26 regions in the whole patient group, at a much greater frequency in the first-episode group (54 regions), compared to the chronic group (20 regions; Chi-square test, $p=9.5\times10^{-8}$).

Table 1. Differences of lnCVR between patients and healthy controls in females and males.

| Group 1 | Group 2 | Number of regions with higher lnCVR in group 1 compared to group 2 | Number of regions with lower lnCVR in group 1 compared to group 2 |
|---|---|---|---|
| Schizophrenia all (N=1792) | HC all (N=1523) | 50 | 0 |
| FES drug-naive all (N=478) | HC all (N=1523) | 73 | 0 |
| Chronic all (N=398) | HC all (N=1523) | 28 | 4 |
| Schizophrenia female (N=760) | HC female (N=698) | 17 | 0 |
| FES drug-naive female (N=239) | HC female (N=698) | 65 | 0 |
| Chronic female (N=139) | HC female (N=698) | 5 | 10 |
| Schizophrenia male(N=1032) | HC male (N=825) | 26 | 0 |
| FES drug-naive male (N=239) | HC male (N=825) | 54 | 0 |
| Chronic male (N=259) | HC male (N=825) | 20 | 3 |

FES, first-episode; HC, healthy control.

#### Result S2. Identifying regions that are ‘invariant’ in gray matter reduction

We derived a score [i.e., rank(ES/CVR)] by ranking ratio between the effect size (ES) in group-level mean difference and variability for each region to identify regions that are ‘invariant’ in gray matter reduction. A small CVR value represents a low variability in gray matter volume (that is, a large 1/CVR represents an ‘invariant’ in gray matter volume). A high ES value represents a large reduction of gray matter volume in patients relative to healthy controls. The table 2 in below shows the top 20 regions with largest ES/CVR value. It shows that the left insula, supramarginal and right prarcentral exhibit ‘invariant’ in gray matter reduction.

Table 2. The top 20 regions with largest ES/CVR value.

| Region | ES/CVR | Rank(ES/CVR) | Rank(ES) | Rank(1/CVR) |
|---|---|---|---|---|
| L_hippocampus | 0.333 | 1 | 1 | 78 |
| R_hippocampus | 0.311 | 2 | 2 | 70 |
| L_insula | 0.282 | 3 | 7 | 12 |
| L_precentral | 0.279 | 4 | 4 | 39 |
| R_thalamus | 0.274 | 5 | 3 | 80 |
| L_superiorfrontal | 0.261 | 6 | 5 | 71 |
| R_fusiform | 0.260 | 7 | 8 | 47 |
| L_postcentral | 0.259 | 8 | 10 | 24 |
| L_superiortemporal | 0.256 | 9 | 9 | 56 |
| R_insula | 0.251 | 10 | 11 | 29 |
| L_thalamus | 0.246 | 11 | 6 | 81 |
| L_supramarginal | 0.241 | 12 | 16 | 10 |
| L_amygdala | 0.231 | 13 | 17 | 31 |
| L_lateraloccipital | 0.231 | 14 | 19 | 25 |
| R_inferiortemporal | 0.229 | 15 | 15 | 48 |
| L_fusiform | 0.228 | 16 | 13 | 67 |
| R_paracentral | 0.227 | 17 | 30 | 4 |
| R_superiorfrontal | 0.226 | 18 | 14 | 63 |
| R_inferiorparietal | 0.226 | 19 | 24 | 30 |
| R_transversetemporal | 0.226 | 20 | 21 | 37 |

#### Result S3. Association between PD- map and human brain gene map.

There was a spatial association (*r*=0.391, *p*=0.022) between the PD- map and gene PLS1 map (see **Supplementary Figure 2**). A total of 1,572 significant genes (FDR *p*<0.05) remained (**Supplementary Figure 2c**), and their enrichment biological processes are shown in the **Supplementary Figure 2d**. In cell type signatures, these genes were enriched in the microglia and astrocytes in the prefrontal cortex, as well as midbrain neuron types (**Supplementary Figure 2e**). In addition, these enriched genes were also related to human brain disorders such as the movement disorders, nystagmus, and hyporeflexia (**Supplementary Figure 2f**).

**Possible interpretation for PD- :** By mapping the proportion of patients with a moderate deviation in GMV reduction, PD- can be hypothesized as a common representation across individuals with schizophrenia, which reflects brain biology involving in ageing, cognitive decline and cortical volume loss. The strongest peak of PD- map was located at bilateral thalamus, indicating that the thalamus is probably one of the most frequently affected regions across all people with schizophrenia. This may be due to thalamic function and connection. Specifically, thalamus plays an important role in integrating sensory input from external environment [20]. It also acts as an interface of body-mind in stress response [21], which is impaired in patients with schizophrenia [22, 23]. Thalamus has widespread connections to different cortical areas, subcortical regions and cerebellum [24]. As a hub, it is suggested to probably be more susceptible to affecting other regions, or be affected. Furthermore, genes associated with PD- were primarily enriched in the astrocytes and microglia in the prefrontal cortex. Interestingly, our finding supports a novel perspective from a recent study reporting that schizophrenia and ageing have a common neurobiological basis, and astrocytes is involved in the multiple kinds of pathophysiology of schizophrenia [25]. These findings suggest that the PD- map can capture certain shared biological signature referring to pathophysiological 'brain ageing' that are most common across people with schizophrenia.

#### Result S4. Association between regional variability in schizophrenia and neurotransmitter systems.

We found significant correlations between brain variability map and neurotransmitter maps (**Supplementary Table 7**). Specifically, the lnCVR map in schizophrenia spatially correlated with the opioid map (MOR, *r*=0.331, *p*=0.036). The PD- map was correlated with neurotransmitter maps in serotonin ($5\text{-}HT_{2a}$, *r*=0.299, *p*=0.006) and dopamine (DAT, *r*=-0.242, *p*=0.022). Additionally, there were significant negative correlations between the PD+ and serotonin $5\text{-}HT_{1A}$, receptors (*r*=-0.296, *p*=0.036) while positive correlations seen with dopamine $D_1$ receptor (*r*=0.337, *p*=0.004).

#### Result S5. Network-based diffusion modeling at group-level and individual-level.

We employed the NDM to simulate the possible processes (i.e., spatial 'spreading' pattern) of GMV alteration across brain regions. We assessed the model performance by using the Spearman correlation coefficient between the spatial map of group-level GMV difference (i.e., effect size map) and the NDM simulated map. We found that the GMV difference in patients was successfully simulated by using the NDM (**Supplementary Figure 2a and Supplementary Table 8**). We observed significant spatial correlations between observed values and simulated values for three models: $NDM_{MC}$ (*r*=0.789, $p=1.0\times10^{-18}$), $NDM_{SC}$ (*r*=0.547, $p=1.1\times10^{-7}$), and $NDM_{FC}$ (*r*=0.669, $p=6.2\times10^{-12}$) (**Supplementary Figure 3a**). Consistent with discovery sample, the spatial correlations were replicated in the validation sample (**Supplementary Figure 3a**). Temporal lobe regions (mainly the hippocampus) were consistently identified as the optimal source seed (**Supplementary Figure 3b**), across the three models, and across subsamples at different illness stages (first-episode, drug-naïve, and chronic) (**Supplementary Table 8**). In the validation data, additional optimal seeds were found in the frontal pars opercularis region by the $NDM_{MC}$. In both subsamples with first-episode and drug-naïve patients, additional optimal seeds were found in the thalamus by the $NDM_{SC}$ (**Supplementary Table 8**). A full list is provided in **Supplementary Table 9** to show the model performances with each region being the starting point of NDM. This indicates that 'spreading' atrophy patterns starting at hippocampus, pars opercularis and thalamus are likely explanations for the variability seen in patients.

At the individual-level, we employed the NDM to estimate the individual deviations in GMV differences in each patient. We found that the NDM can achieve an accepted performance ($p_{spin}$<0.001) in the consistency between estimated values and observed values for 72.1% patients ($SZ_{NDM}$, n=1,292) (**Supplementary Table 10a**). For these $SZ_{NDM}$ patients, we further classified them into several subgroups based on the heterogeneity in optimal seed locations. The seed of NDM may serve as the 'disease epicenters' that lead to network-spreading patterns of brain structural changes during regional pathological processes [13]. The heterogeneity of pathological processes may be uncovered by detecting the optimal seeds applying the NDM on individual patient's data. Herein, patients were assigned to subgroups, termed as 'spatial' phenotypes, based on the spatial locations of the seeds (**Supplementary Table 10b**). **Figure 4** shows specific spatial patterns of GMV change in each spatial phenotype, which are labelled as 'temporal cortex', 'frontal cortex', 'occipital cortex', 'parietal cortex', 'somatosensory cortex', 'cingulate-insula', 'basal ganglia' or 'thalamus' phenotypes. It shows that 'temporal cortex' and 'frontal cortex' phenotypes account for the highest proportion of patients, reaching 24.2% and 13.6% respectively. **Supplementary Table 11** shows regional comparison of GMV between patients and controls for each spatial phenotype. Together, we show the existence of neuroanatomically distinct phenotypes, which are characterized with differential spatial patterns of gray matter alterations in schizophrenia.

#### Discussion S1. Lower variability in schizophrenia is inconsistent in prior studies.

The regional distribution of lower variability in schizophrenia (i.e., homogeneity) has been inconsistent in prior studies [26-28]. Previous study reported a lower variability in the anterior cingulate volume [26] but this was not replicated in two other recent works [27, 28]. In contrast, a recent work found significantly lower variability in the left parahippocampal region [27]. Our data did not show reduced variability in any regional volume in first-episode patients, but in established cases, variability is lower in the left parahippocampal and specific occipital regional volumes. Notably, occipital regions exhibit higher volume, indicating that over time some shared tissue increase may occur among patients, irrespective of the heterogenous origins of the tissue reduction, supporting a dynamic reorganization effect [29]. Importantly, we find that higher inter-individual variability is associated with a reduction in GMV, but not with the increase seen across the brain, supporting the notion that compensatory activity may occur over time [30]. Taken together, our work provides a comprehensively evaluation on the inter-patient variability of structural brain measures, summarizing consistent/inconsistent findings in heterogeneity and homogeneity of neuroanatomical alteration in schizophrenia, compared with previous knowledge [26-28].

**Supplementary Table 1. Sample demographics by participating cohort.**

| Cohort | N | | Mean Age (SZ) | Mean Age (HC) | F/M (SZ) | F/M (HC) | Mean Duration Of Illness (years) | PANSS Positive Subscale | PANSS Negative Subscale | PANSS General Subscale | PANSS Total Score |
|---|---|---|---|---|---|---|---|---|---|---|---|
| | Case | Control | | | | | | | | | |
| CN-Shanghai1 | 361 | 267 | 24.3±8.0 | 24.0±4.9 | 167/194 | 144/123 | 0.9±1.4 | 20.2±5.7 | 15.4±7.4 | 38.1±8.1 | 73.7±15.7 |
| CN-Shanghai2 | 159 | 133 | 24.9±7.0 | 23.7±5.9 | 84/75 | 67/66 | 1.4±2.0 | 23.7±5.3 | 18.1±6.8 | 41.4±7.2 | 83.2±13.7 |
| CN-Shanghai3 | 42 | 23 | 29.9±7.4 | 31.2±5.9 | 23/19 | 12/11 | 6.6±5.7 | 19.9±3.1 | 18.4±6.4 | 33.1±4.9 | 71.4±9.0 |
| CN-Harbin | 49 | 0 | 26.7±8.1 | - | 22/27 | - | 2.4±3.2 | 20.2±4.5 | 22.4±6.4 | 40.4±7.4 | 83.0±14.5 |
| CN-Chengdu | 100 | 127 | 40.2±11.8 | 38.1±14.9 | 30/70 | 42/85 | 15.5±10.3 | 13.2±5.8 | 21.0±6.3 | 28.2±5.7 | 62.3±13.1 |
| CN-Taibei | 158 | 254 | 43.9±11.0 | 38.0±12.0 | 93/65 | 136/118 | 15.7±10.1 | 9.6±3.2 | 10.0±5.5 | 20.7±4.4 | 40.3±10.7 |
| CN-Zhengzhou | 194 | 59 | 23.2±8.6 | 26.5±5.5 | 97/97 | 32/27 | - | 22.3±5.1 | 23.1±6.3 | 44.1±7.9 | 89.5±15.0 |
| CN-Beijing | 130 | 0 | 21.3±4.2 | - | 75/55 | - | - | 22.4±5.0 | 20.9±7.2 | 38.8±7.0 | 82.1±15.1 |
| CN-Xian | 39 | 51 | 23.9±8.6 | 21.1±4.0 | 17/22 | 18/33 | 1.2±1.8 | 22.9±4.9 | 21.4±6.4 | 44.8±8.8 | 89.1±14.6 |
| HCP-EP | 52 | 39 | 21.9±3.0 | 25.4±4.4 | 15/39 | 14/25 | - | 11.7±3.9 | 16.0±5.6 | 24.9±4.5 | 52.7±10.1 |
| COBRE | 79 | 86 | 37.8±13.8 | 38.7±11.9 | 15/64 | 23/63 | 15.6±12.6 | 15.2±5.0 | 15.2±5.4 | 29.2±9.1 | 59.6±15.6 |
| fBIRN | 42 | 65 | 38.1±11.8 | 38.3±11.2 | 14/28 | 29/36 | - | - | - | - | - |
| MCIC | 109 | 95 | 33.8±11.2 | 32.9±12.2 | 26/83 | 30/65 | 11.1±10.9 | - | - | - | - |
| NMorphCH | 44 | 43 | 32.5±6.9 | 31.5±8.4 | 14/30 | 22/21 | - | - | - | - | - |
| NUSDAST | 123 | 127 | 33.8±12.5 | 32.6±13.8 | 42/81 | 62/65 | - | - | - | - | - |
| DS000030 | 50 | 121 | 36.5±8.9 | 31.6±8.8 | 12/38 | 56/65 | - | - | - | - | - |
| DS000115 | 22 | 18 | 24.2±3.8 | 20.7±4.8 | 6/16 | 8/10 | - | - | - | - | - |
| DS004302 | 46 | 24 | 42.5±10.7 | 39.5±14.3 | 10/36 | 7/17 | - | - | - | - | - |
| Total | 1799 | 1532 | 29.9±11.9 | 31.3±11.8 | 762/1037 | 702/830 | 7.4±10.0 | 18.7±6.9 | 17.3±7.6 | 35.4±10.5 | 71.3±20.7 |

**Supplementary Table 2. Dataset-specific information.**

| Cohort | Diagnosis measurement | Exclusion/inclusion criteria | Scanner manufacturer and type | Imaging protocols |
|---|---|---|---|---|
| CN-Shanghai1 | DSM-IV | All the subjects were Mandarin-speaking Han Chinese individuals from Shanghai metropolitan area. The FES patients were identified according to DSM-IV criteria by qualified psychiatrists using all available clinical information including a diagnostic interview of patients and their family, clinical case notes, and clinician's observations. All healthy subjects were assessed in accordance with DSM-IV criteria as being free of schizophrenia and other axis I disorders, and none had neurological diseases, head trauma, or substance abuse. All participants were right-handed; had no history of substance abuse or suicidal ideation; and had no MRI contraindications. | GE Sigma 3T | GE Sigma 3.0 T MR (GE Medical Systems, Milwaukee, Wisconsin). TR = 7.8 ms; TE = 3.65 ms; flip angle = 7°; matrix = 256×256; voxel size = 1×1×1 mm. |
| CN-Shanghai2 | DSM-IV | The exclusion criteria were as follows: (1) brain trauma, substance-related disorders, major medical or neurologic disorders; (2) other mental disorders meeting DSM-IV criteria; (3) drug or alcohol abuse; (4) pregnancy, breastfeeding or other unstable clinical state including aggressive behaviour; (5) history of electroconvulsive therapy or transcranial magnetic stimulation within six months; and (6) other contraindications to MRI scanning. To eliminate potential familial effects, healthy subjects whose first- or second-degree relatives had a history of mental disorders were also excluded. | 3T Siemens MR B17 | A 3-Tesla MRI scanner (Siemens MR B17) was used to acquire data in the Shanghai Mental Health Centre. High-spatial-resolution T1-weighted images were collected by a magnetization-prepared rapid acquisition gradient echo (MPRAGE) sequence. The main parameters included repetition time/echo time, 2530/2.56 msec; flip angle, 7°; field of view, 256 × 256 mm2; matrix size, 256 × 256; section thickness, 1 mm (no gap); and voxel size, 1×1×1mm3. |
| CN-Shanghai3 | DSM-IV-TR | Inpatients with schizophrenia were recruited from the Shanghai Mental Health Center (SMHC). All the patients were recruited from October 2013 to January 2015. All patients met criteria for schizophrenia or schizoaffective disorder based on the Structured Clinical Interview for Diagnostic and Statistical Manual of Mental Disorders (DSM-IVTR) as conducted by senior psychiatrists. The severity of psychotic symptoms was assessed by the Positive and Negative Syndrome Scale (PANSS). All patients had a total PANSS score of 60 or more and had not received ECT during the past six months. All health controls did not have a lifetime psychiatric disorder or family history of psychosis in their first-degree relatives. Participants were excluded if they had brain injuries, organic mental disorders, neurologic abnormalities, other serious physical illnesses, dementia, substance abuse or dependence, or contraindications to MRI. | 3-T Siemens Magnetom Verio Syngo MR B17 | High-resolution T1-weighted images were acquired using a 3-T Siemens Magnetom Verio Syngo MR B17 scanner. High-resolution T1-weighted images were collected with a magnetization-prepared rapid acquisition gradient echo (MPRAGE) sequence (TR = 2530 ms; TE = 2.56 ms; flip angle = 7°; inversion time = 1100 ms; FOV = 256 mm × 256 mm; matrix = 256 × 256; slice thickness = 1 mm; 224 slices; and voxel size = 1.0 × 1.0 × 1.0 mm). |
| CN-Harbin | DCM-IV SCID | Patient recruitment criterion was based on the Diagnostic and Statistical Manual of Mental Disorders, Fourth Edition and Structured Clinical Interview for DSM (SCID), which includes: (1) Age range between 14-45 years old; (2) Intelligence quotient (IQ)>69; (3) The first onset and no systematic antipsychotic treatment before admission; (4) Psychotic symptom assessment scale (positive and negative symptom scale score) is greater than or equal to 60; clinical overall | 3.0 Tesla GE Discovery MR750 | All scans were acquired on the same 3.0 Tesla GE Discovery MR750 scanner equipped with a 32-channel head coil. |

| | | | | |
|---|---|---|---|---|
| | | impression CGI severity is greater than or equal to 4 points; (5) Exclude other Axis I mental disorders except schizophrenia, and have no DSM-5 drug or alcohol dependence within the past three months. Exclusion criteria for patients included the following: (1) Sensory-motor disturbances (hearing impairment, blindness), neurological disorders (brain injury, epilepsy), or other medical conditions (2) claustrophobic and unable to undergo MRI scans; (3) patients with repetitive transcranial magnetic stimulation or MRI Contraindications to scanning, such as those with metal implants in the body; (4) suicide attempters, pregnant or breastfeeding women. | | |
| CN-Chengdu | DSM-IV | Patients with SZ were recruited from the Clinical Hospital of Chengdu Brain Science Institute. Each patient was diagnosed based on the Diagnostic and Statistical Manual of Mental Disorders, Fourth Edition (DSM-IV). Subjects with a history of brain injuries, substance-related disorders, major medical or neurological disorder were excluded. To exclude the potential effect of similar genetic backgrounds, the history of psychiatric disorder in a first- or second-degree relative was an additional exclusion criterion for healthy controls. | 3-Tesla GE DISCOVERY MR 750 | A 3-Tesla MRI scanner (GE DISCOVERY MR 750, USA) was used to collect imaging data in the University of Electronic Science and Technology of China. High-resolution T1-weighted images were acquired using a three dimensional fast spoiled gradient echo (T1-3D FSPGR) sequence. The main parameters include: TR = 6.008 ms; TE = 1.984 ms; flip angle (FA) =90°; field of view (FOV) = 25.6 cm × 25.6 cm; matrix size = 256 × 256; slice thickness = 1 mm (no gap). |
| CN-Taibei | DSM-IV | Patients and matched healthy controls were recruited from the Veteran General Hospital in Taipei, Taiwan. All participants were diagnosed according to the Diagnostic and Statistical Manual of Mental Disorder-IV criteria for schizophrenia, and each participant's history of medical disease, psychiatric illness, and medication use was evaluated by interview and medical charts carefully. Any participants with the following conditions were excluded: (1) a comorbid substance-related disorder, (2) presence of neurobiological disorders, such as dementia, head injury, stroke, or Parkinson’s disease; (3) presence of hypertension, diabetes, hyperlipidemia or coronary heart disease; (4) severe medical illness, such as malignancy, heart failure, or renal failure; (4) presence of ferromagnetic foreign bodies or implants that were anywhere in the body. | 3T Siemens | Siemens MAGNETOM Tim Trio 3.0T MRI Scanner (Siemens Healthcare, Erlangen, Germany) with a 12- channel head coil, using 3-dimensional magnetization prepared rapid gradient-echo sequence. TR: 2530 ms, TE: 3.5 ms, TI: 1,100 ms, FoV: 256 mm, flip angle: 7 degree, 192 sagittal slices, voxel size = 1.0 mm3 cubic, no gap. |
| CN-Zhengzhou | DSM-IV | All patients were identified according to the Diagnosis and Statistic Manual of Mental Disorders, fourth edition (DSM-IV) criteria for schizophrenia by qualified psychiatrists using all available clinical information including a diagnostic interview, clinical case notes, and clinician’s observations. The severity of positive and negative symptoms was assessed by trained and experienced psychiatrists using PANSS scoring. Individuals were excluded from the study if they were diagnosed with schizoaffective disorder, mood disorders, other cognitive disorders, epilepsy, had severe physical diseases, alcohol/drug dependence, or been treated with electroconvulsive therapy. Healthy subjects were assessed in accordance with DSM-IV criteria as being free of schizophrenia and other Axis I disorder, and none had neurological diseases, head trauma, substance abuse, suicidal ideation, and MRI contraindications. | GE MR750 3T | GE MR750 3T MRI scanner with standard quadrature head coil. TR/TE= 8.2 / 3.2 ms, FOV= 256×256 mm 2, Matrix= 256×256, slice thickness= 1.0 mm, gap= 0mm, 188 contiguous sagittal slices；flip angle = 12°. |

| | | | | |
|---|---|---|---|---|
| CN-Beijing | DSM-IV | The inclusion criteria were: (1) both in- and out-patients, (2) both genders, (3) diagnosis of schizophrenia established with Structured Clinical Interview for the DSM-IV Axis I Disorder, patient edition, (4) aged between 18 and 45 years, (5) onset at age ≥ 15 years, (6) first episode of schizophrenia, (7) no previous psychiatric treatment ('drug-naïve' status), and (8) ability to understand the contents of interview and provide written informed consent.<br>The exclusion criteria were: (1) a history or diagnosis of major medical conditions, (2) a history of alcohol and / or drug abuse or dependence, and (3) contra-indication to olanzapine, aripiprazole, or risperidone. | Siemens Trio Trim 3.0 T | Main parameters: TR: 9.8 ms, TE: 3.8 ms, TI: 450 ms, FoV: 512 x 512, flip angle: 13, slice thickness=1.2 mm. |
| CN-Xian | DSM-IV, DSM-V | Inclusion criteria were the following: (1) diagnosis of schizophrenia using the Structured Clinical Interview for the Diagnostic and Statistical Manual of Mental Disorders, Fourth Edition (SCID-IV), (2) patients were taking stable doses of antipsychotic medication for at least 8 weeks before the study; (3) no history of significant head trauma or neurological disorders and no focal brain lesions by T1- or T2-weighted MRI; (4) no alcohol or drug abuse; and (5) patient has no recent aggression or other forms of behavioral dysfunction. | GE Discovery MR750 3.0 T | High-resolution T1-weighted MRI was acquired using a GE Discovery MR750 3.0 T scanner located in the Department of Radiology, and all subjects underwent T2WI scans to rule out organic diseases. The scanning parameters were repetition time 8.2 ms, echo time 3.2 ms, flip angle 12°, field of view 256 × 256 mm, matrix 256 × 256, slice thickness 1 mm, and sagittal slices 196. |
| HCP-EP | DSM-V | Available at https://www.humanconnectome.org/study/human-connectome-project-for-early-psychosis | | |
| COBRE | SCID DSM-IV Axis I Disorders | All participants were in the 18-65 age range and had a diagnosis of schizophrenia. Healthy individuals were included if they did not have a personal or family history of psychiatric disorders. History of neurological disorder, history of mental retardation, history of severe head trauma with more than 5 minutes loss of consciousness, history of substance abuse or dependence within the last 12 months and MRI contraindications. | 3T Siemens TIM Trio | T1-weighted images were acquired with a 5-echo multi-echo MPRAGE sequence [TE (echo times) = 1.64, 3.5, 5.36, 7.22, 9.08 ms, TR (repetition time) = 2.53 s, TI (inversion time) = 1.2 s, 7° flip angle, number of excitations (NEX) = 1, slice thickness = 1 mm, FOV (field of view) = 256 mm, resolution = 256x256]. |
| fBIRN | SCID DSM-IV-TR | All subjects diagnosed with schizophrenia were clinically stable outpatients whose antipsychotic medications and doses had not changed within the last two months. Schizophrenia and healthy volunteers with a history of major medical illness, drug dependence in the last five years (except for nicotine), current substance abuse disorder, or MRI contraindications, were excluded. Individuals with schizophrenia who had significant tardive dyskinesia and healthy volunteers with a current or past history of major neurological or psychiatric illness or with a first-degree relative with a DSM IV Axis-I psychotic disorder diagnosis were also excluded. | 3T Siemens Tim Trio; 3T GE | High-resolution structural imaging scans were acquired on six 3T Siemens Tim® Trio System and one 3T General Electric Discovery MR750 scanner. MP-RAGE scan parameters for the Siemens scanner were: scan plane=sagittal, TR/TE/TI=2300/2.94/1100ms, GRAPPA acceleration factor=2, flip angle=9°, resolution=256×256x160, FOV=220mm2, voxel size=0.86x0.86x1.2mm, and NEX=1. IR-SPGR scan parameters for the General Electric scanner were: scan plane=sagittal, TR/TE/TI=5.95/1.99/450ms, ASSET acceleration factor=2, a flip angle=12°, resolution=256×256x166, FOV=220mm2, voxel size=0.86x0.86x1.2mm, and NEX=1. All scans covered the entire brain. |
| MCIC | SCID DSM-IV (SCID-NP for controls) or CASH | All subjects were between the ages of 18 and 60 and spoke English as their native language. To be included in the schizophrenia cohort, patients had to meet diagnostic criteria for schizophrenia, schizoaffective disorder, or schizophreniform disorder. Concerted effort was made to recruit patients | 1.5, 3T Siemens and GE | T1 scans: TR = 2530 ms for 3 T, TR = 12 ms for 1.5 T; TE = 3.79 ms for 3 T, TE = 4.76 ms for 1.5 T; FA = 7 for 3 T, FA = 20 for 1.5 T; TI = 1100 for 3 T; Bandwidth = 181 for 3 T, Bandwidth = 110 for 1.5 T; 0.625×0.625 mm voxel |

| | | | | |
|---|---|---|---|---|
| | were used to diagnose primary and co-morbid psychiatric disorders in controls and patients | early in the course of their illness and especially those who were antipsychotic drug naïve. The healthy control subjects with no current or past history of psychiatric illness including substance abuse or dependence were matched within site to the patient cohort for age, sex, and parental education. Control subjects who had not been diagnosed with any psychiatric disorders, but had been medicated with antidepressants, anti-anxiety medication or medication for sleep disturbance were included in the study provided that the duration of their medication did not exceed 2 months of lifetime use and no medication was used within the 6 months preceding the baseline MRI scan. Control subjects who met criteria for current or past history of substance abuse or dependence were excluded from the study. Patients, however, were not excluded from the study unless criteria were met for current (i.e., within the past month) abuse or dependence (except for 6 patients who were found to meet criteria for current abuse after the study data was collected). Both patients and controls were excluded if they had (1) an IQ less than 70 based on a standardized IQ test, (2) history of a head injury resulting in prolonged loss of consciousness, neurosurgical procedure, neurological disease, history of skull fracture, severe or disabling medical conditions, or (3) a contraindication for MRI scanning such as pregnancy, metal in body or head including implanted pacemaker, medication pump, vagal stimulator, deep brain stimulator, implanted TENS unit, or ventriculo-peritoneal shunt | | size; slice thickness 1.5 mm; FOV 256×256×128 cm matrix; FOV = 16 cm (could be increased to 18 cm when needed for full brain coverage). |
| NMorphCH | This is a longitudinal study examining the clinical, cognitive and neuroimaging (MRI) data from schizophrenia and control subjects at baseline and after two years. The study was conducted at Northwestern University. Neuroimaging data includes T1, T2, DTI, resting-state fMRI and n-back task fMRI. More details are provide at http://www.schizconnect.org/documentation | | 3T | MPRAGE:voxel size =1x1x1.6 mm3; Matrix size 256x256; TR=3.15ms; TE=20ms; Flip ange=8.0 |
| NUSDAST | The Northwestern University Schizophrenia Data and Software Tool (NUSDAST) is a repository of schizophrenia neuroimaging data collected from over 450 individuals with schizophrenia, healthy controls and their respective siblings, most with 2-year longitudinal follow-up. More details are provided at: Wang L, Kogan A, Cobia D, et al. Northwestern University schizophrenia data and software tool (NUSDAST). Frontiers in neuroinformatics, 2013, 7: 25. | | Siemens 1.5 T Vision scanner | 3D MPRAGE (TR = 9.7 ms, TE = 4 ms, flip = 10°, ACQ = 1, 256 × 256 matrix, 1 × l mm in-plane resolution, 128 slices, slice thickness 1.25 mm) |
| DS000030 | DSM-IV | For both healthy and patient groups, participants were men or women ages 21–50 years; NIH racial/ethnic category either White, not Hispanic or Latino; or Hispanic or Latino, of any racial group; primary language either English or Spanish; completed at least 8 years of formal education; no significant medical illness; adequate cooperation to complete assessments; visual acuity 20/60 or better; and urinalysis negative for drugs of abuse (Cocaine; Methamphetamine; Morphine; THC; and Benzodiazepines).<br>Participants in the healthy group were excluded if they had lifetime diagnoses of Schizophrenia or Other Psychotic Disorder, Bipolar I or II Disorder, or Substance Abuse or Dependence (not counting caffeine or nicotine); or current Major Depressive Disorder; suicidality; Anxiety Disorder | Siemens Trio (2 Imaging Sites) | A T1-weighted high-resolution anatomical scan (MPRAGE) were collected with the following parameter: slice thickness = 1mm, 176 slices, TR=1.9s, TE=2.26ms, matrix=256 x 256, FOV=250mm. |

| | | | | |
|---|---|---|---|---|
| | | (Obsessive Compulsive Disorder, Panic Disorder, Generalized Anxiety Disorder, Post-Traumatic Stress Disorder), Attention Deficit Hyperactivity Disorder (ADHD). ADHD criteria were assessed using Adult ADHD Interview; healthy participants were screened for sub-threshold ADHD, defined as 4 or more ADHD inattentive or hyperactive/impulsive symptoms in either childhood and adulthood; in addition, they could not have had medication treatment for ADHD within the prior 12 months. Each of the patient groups (Schizophrenia, Bipolar Disorder, and ADHD) excluded anyone with one of these other diagnoses; stable medications were permitted for the patients. For MRI studies we excluded participants who were left handed, who believed they might be pregnant, or had other contraindications to scanning (e.g., claustrophobia, metal in body, body too large to fit in scanner).<br>More details are provided at https://www.nature.com/articles/sdata2016110 | | |
| DS000115 | DSM-IV | All subjects were diagnosed on the basis of a consensus between a research psychiatrist who conducted a semi-structured interview and a trained research assistant who used the Structured Clinical Interview for DSM-IV Axis I Disorders. Participants were excluded if they: (a) met DSM-IV criteria for substance dependence or severe/moderate abuse during the prior 6 months; (b) had a clinically unstable or severe medical disorder; (c) had a history of head injury with documented neurological sequelae or loss of consciousness; or (d) met DSM-IV criteria for mental retardation. The individuals with schizophrenia were all outpatients, and were stabilized on antipsychotic medication for at least 2 weeks. Controls were required to have no lifetime history of Axis I psychotic or mood disorders and no first-degree relatives with a psychotic disorder. | 3T Siemens Trio | T1 structural image was acquired using a sagittal MP-RAGE 3D sequence (TR = 2400 ms, TE = 3.16 ms, flip = 8°; voxel size = 1 mm × 1 mm × 1 mm). |
| DS004302 | DSM-V | Patients meeting DSM-5 criteria for schizophrenia were initially recruited from four psychiatric hospitals in Barcelona. Diagnoses were made using the Structured Clinical Interview for DSM Disorders (SCID). Patients were excluded if they (a) were younger than 18 or older than 65, (b) had a history of brain trauma or neurological disease or (c) had shown alcohol/substance abuse/dependence within 12 months prior to participation. Social use of alcohol was permitted, as was non-habitual use of cannabis. Electroconvulsive therapy in the past 6 months was also an exclusion criterion. All participants were right-handed and were taking antipsychotic medication. Healthy controls met the same exclusion criteria as the patients, and they were also interviewed using the SCID to exclude current and past psychiatric disorders. They were questioned and excluded if they reported a history of treatment with psychotropic medication beyond non-habitual use of night sedation. Controls were also excluded if they reported a history of major psychiatric disorder in a first-degree relative. | 3T Philips Ingenia scanner | Images were acquired with a 3T Philips Ingenia scanner (Philips Medical Systems, Best, The Netherlands). High-resolution anatomical volume with an FFE (Fast Field Echo) sequence for anatomical reference and inspection (TR = 9.90ms; TE = 4.60ms; Flip angle = 8°; voxel size = 1 × 1mm; slice thickness = 1mm; slice number = 180; FOV = 240mm). |

**Supplementary Table S3. Mean difference in regional gray matter volume measures**

| Classification | Region | Effect size (Cohend's d) | t test | | |
|---|---|---|---|---|---|
| | | | t value | p value | FDR(p<0.05) |
| Thalamus | L_thalamus | 0.29 | -8.29 | 1.11E-16 | * |
| Basal Ganglia | L_caudate | -0.05 | 1.65 | 9.93E-02 | |
| Basal Ganglia | L_putamen | -0.04 | 0.84 | 4.00E-01 | |
| Basal Ganglia | L_pallidum | -0.13 | 4.10 | 4.31E-05 | * |
| Hippcampus | L_hippocampus | 0.38 | -12.00 | 0.00E+00 | * |
| Amygdala | L_amygdala | 0.25 | -7.55 | 5.55E-14 | * |
| Basal Ganglia | L_accumbens | 0.21 | -6.10 | 1.20E-09 | * |
| Thalamus | R_thalamus | 0.32 | -9.17 | 0.00E+00 | * |
| Basal Ganglia | R_caudate | -0.07 | 2.18 | 2.96E-02 | |
| Basal Ganglia | R_putamen | -0.06 | 1.55 | 1.21E-01 | |
| Basal Ganglia | R_pallidum | -0.16 | 4.86 | 1.21E-06 | * |
| Hippcampus | R_hippocampus | 0.35 | -10.69 | 0.00E+00 | * |
| Amygdala | R_amygdala | 0.16 | -5.16 | 2.68E-07 | * |
| Basal Ganglia | R_accumbens | 0.24 | -7.18 | 8.50E-13 | * |
| Temporal | L_bankssts | 0.14 | -3.74 | 1.86E-04 | * |
| Cingulate | L_caudalanteriorcingulate | 0.11 | -3.01 | 2.62E-03 | * |
| Frontal | L_caudalmiddlefrontal | 0.15 | -4.62 | 4.06E-06 | * |
| Occipital | L_cuneus | 0.12 | -3.34 | 8.38E-04 | * |
| Temporal | L_entorhinal | 0.09 | -2.22 | 2.67E-02 | * |
| Temporal | L_fusiform | 0.26 | -7.80 | 8.55E-15 | * |
| Parietal | L_inferiorparietal | 0.21 | -6.26 | 4.37E-10 | * |
| Temporal | L_inferiortemporal | 0.24 | -7.10 | 1.54E-12 | * |
| Cingulate | L_isthmuscingulate | 0.11 | -3.39 | 7.16E-04 | * |
| Occipital | L_lateraloccipital | 0.25 | -7.01 | 2.89E-12 | * |
| Frontal | L_lateralorbitofrontal | 0.24 | -7.16 | 9.73E-13 | * |
| Occipital | L_lingual | 0.19 | -5.52 | 3.65E-08 | * |
| Frontal | L_medialorbitofrontal | 0.12 | -3.75 | 1.82E-04 | * |
| Temporal | L_middletemporal | 0.23 | -6.72 | 2.09E-11 | * |
| Temporal | L_parahippocampal | 0.16 | -4.75 | 2.14E-06 | * |
| Somatosensory | L_paracentral | 0.17 | -4.84 | 1.37E-06 | * |
| Frontal | L_parsopercularis | 0.18 | -5.41 | 6.74E-08 | * |
| Frontal | L_parsorbitalis | 0.17 | -5.22 | 1.87E-07 | * |
| Frontal | L_parstriangularis | 0.16 | -4.74 | 2.27E-06 | * |
| Occipital | L_pericalcarine | 0.04 | -0.74 | 4.58E-01 | |
| Somatosensory | L_postcentral | 0.27 | -7.95 | 2.55E-15 | * |
| Cingulate | L_posteriorcingulate | 0.18 | -5.79 | 7.70E-09 | * |
| Somatosensory | L_precentral | 0.30 | -8.74 | 0.00E+00 | * |
| Parietal | L_precuneus | 0.18 | -5.41 | 6.67E-08 | * |
| Cingulate | L_rostralanteriorcingulate | 0.16 | -4.33 | 1.52E-05 | * |
| Frontal | L_rostralmiddlefrontal | 0.19 | -5.83 | 6.10E-09 | * |
| Frontal | L_superiorfrontal | 0.30 | -8.75 | 0.00E+00 | * |
| Parietal | L_superiorparietal | 0.15 | -4.41 | 1.07E-05 | * |
| Temporal | L_superiortemporal | 0.28 | -8.23 | 2.22E-16 | * |
| Parietal | L_supramarginal | 0.25 | -7.10 | 1.56E-12 | * |
| Frontal | L_frontalpole | 0.08 | -2.61 | 9.10E-03 | * |
| Temporal | L_temporalpole | 0.03 | -0.76 | 4.45E-01 | |

| | | | | | |
|---|---|---|---|---|---|
| Temporal | L_transversetemporal | 0.17 | -5.20 | 2.10E-07 | * |
| Insula | L_insula | 0.29 | -8.70 | 0.00E+00 | * |
| Temporal | R_bankssts | 0.18 | -5.31 | 1.16E-07 | * |
| Cingulate | R_caudalanteriorcingulate | 0.11 | -3.09 | 2.05E-03 | * |
| Frontal | R_caudalmiddlefrontal | 0.15 | -4.30 | 1.78E-05 | * |
| Occipital | R_cuneus | 0.16 | -4.57 | 5.04E-06 | * |
| Temporal | R_entorhinal | 0.07 | -1.95 | 5.18E-02 | |
| Temporal | R_fusiform | 0.28 | -8.65 | 0.00E+00 | * |
| Parietal | R_inferiorparietal | 0.24 | -7.06 | 2.05E-12 | * |
| Temporal | R_inferiortemporal | 0.25 | -7.35 | 2.57E-13 | * |
| Cingulate | R_isthmuscingulate | 0.14 | -4.65 | 3.38E-06 | * |
| Occipital | R_lateraloccipital | 0.24 | -6.95 | 4.30E-12 | * |
| Frontal | R_lateralorbitofrontal | 0.20 | -5.69 | 1.38E-08 | * |
| Occipital | R_lingual | 0.17 | -5.02 | 5.39E-07 | * |
| Frontal | R_medialorbitofrontal | 0.25 | -7.21 | 7.13E-13 | * |
| Temporal | R_middletemporal | 0.24 | -7.18 | 8.78E-13 | * |
| Temporal | R_parahippocampal | 0.19 | -5.76 | 8.92E-09 | * |
| Somatosensory | R_paracentral | 0.23 | -6.64 | 3.62E-11 | * |
| Frontal | R_parsopercularis | 0.17 | -5.17 | 2.50E-07 | * |
| Frontal | R_parsorbitalis | 0.14 | -4.08 | 4.67E-05 | * |
| Frontal | R_parstriangularis | 0.13 | -3.97 | 7.41E-05 | * |
| Occipital | R_pericalcarine | 0.02 | -0.43 | 6.69E-01 | |
| Somatosensory | R_postcentral | 0.22 | -6.36 | 2.26E-10 | * |
| Cingulate | R_posteriorcingulate | 0.24 | -7.35 | 2.45E-13 | * |
| Somatosensory | R_precentral | 0.23 | -6.87 | 7.45E-12 | * |
| Parietal | R_precuneus | 0.20 | -6.01 | 2.05E-09 | * |
| Cingulate | R_rostralanteriorcingulate | 0.18 | -4.90 | 1.00E-06 | * |
| Frontal | R_rostralmiddlefrontal | 0.15 | -4.49 | 7.30E-06 | * |
| Frontal | R_superiorfrontal | 0.26 | -7.62 | 3.31E-14 | * |
| Parietal | R_superiorparietal | 0.18 | -5.20 | 2.10E-07 | * |
| Temporal | R_superiortemporal | 0.26 | -7.88 | 4.55E-15 | * |
| Parietal | R_supramarginal | 0.20 | -6.01 | 2.10E-09 | * |
| Frontal | R_frontalpole | 0.12 | -3.83 | 1.30E-04 | * |
| Temporal | R_temporalpole | 0.02 | -0.73 | 4.67E-01 | |
| Temporal | R_transversetemporal | 0.24 | -7.16 | 9.77E-13 | * |
| Insula | R_insula | 0.27 | -8.05 | 1.11E-15 | * |

**Supplementary Table S4. Variability difference in regional gray matter volume measures**

| Classification | Region | All patients group | | | First-episode drug-naive group | | | Chronic medicated group | | |
|---|---|---|---|---|---|---|---|---|---|---|
| | | lnCVR | p value | FDR (p<0.05) | lnCVR | p values | FDR (p<0.05) | lnCVR | p values | FDR (p<0.05) |
| Thalamus | L_thalamus | 0.167 | 0.00E+00 | * | 0.285 | 0.00E+00 | * | 0.158 | 0.00E+00 | * |
| Basal Ganglia | L_caudate | 0.135 | 0.00E+00 | * | 0.179 | 0.00E+00 | * | 0.214 | 0.00E+00 | * |
| Basal Ganglia | L_putamen | 0.089 | 2.00E-03 | * | 0.226 | 0.00E+00 | * | 0.088 | 3.20E-02 | |
| Basal Ganglia | L_pallidum | 0.051 | 8.40E-02 | | -0.006 | 8.60E-01 | | 0.091 | 1.50E-02 | * |
| Hippcampus | L_hippocampus | 0.145 | 0.00E+00 | * | 0.176 | 0.00E+00 | * | 0.238 | 0.00E+00 | * |
| Amygdala | L_amygdala | 0.065 | 2.50E-02 | | 0.016 | 6.81E-01 | | 0.070 | 5.50E-02 | |
| Basal Ganglia | L_accumbens | 0.128 | 0.00E+00 | * | 0.162 | 0.00E+00 | * | 0.113 | 0.00E+00 | * |
| Thalamus | R_thalamus | 0.165 | 0.00E+00 | * | 0.289 | 0.00E+00 | * | 0.117 | 1.00E-03 | * |
| Basal Ganglia | R_caudate | 0.143 | 0.00E+00 | * | 0.201 | 0.00E+00 | * | 0.205 | 0.00E+00 | * |
| Basal Ganglia | R_putamen | 0.085 | 9.00E-03 | * | 0.201 | 0.00E+00 | * | 0.115 | 4.00E-03 | * |
| Basal Ganglia | R_pallidum | 0.070 | 1.70E-02 | * | 0.074 | 4.50E-02 | | 0.139 | 1.00E-03 | * |
| Hippcampus | R_hippocampus | 0.127 | 1.00E-03 | * | 0.091 | 1.40E-02 | * | 0.233 | 0.00E+00 | * |
| Amygdala | R_amygdala | 0.067 | 2.50E-02 | | 0.071 | 9.70E-02 | | 0.101 | 1.00E-02 | * |
| Basal Ganglia | R_accumbens | 0.084 | 2.00E-03 | * | 0.065 | 5.10E-02 | | 0.150 | 0.00E+00 | * |
| Temporal | L_bankssts | 0.033 | 2.34E-01 | | 0.107 | 3.00E-03 | * | 0.009 | 8.14E-01 | |
| Cingulate | L_caudalanteriorcingulate | 0.017 | 5.57E-01 | | 0.086 | 1.40E-02 | * | -0.066 | 5.30E-02 | |
| Frontal | L_caudalmiddlefrontal | 0.080 | 4.00E-03 | * | 0.218 | 0.00E+00 | * | 0.073 | 3.00E-02 | |
| Occipital | L_cuneus | -0.002 | 9.24E-01 | | -0.035 | 2.81E-01 | | -0.104 | 1.00E-03 | * |
| Temporal | L_entorhinal | 0.071 | 3.90E-02 | | 0.149 | 0.00E+00 | * | -0.060 | 1.26E-01 | |
| Temporal | L_fusiform | 0.125 | 0.00E+00 | * | 0.179 | 0.00E+00 | * | 0.106 | 3.00E-03 | * |
| Parietal | L_inferiorparietal | 0.061 | 1.70E-02 | * | 0.158 | 0.00E+00 | * | 0.028 | 4.18E-01 | |
| Temporal | L_inferiortemporal | 0.089 | 1.00E-03 | * | 0.132 | 0.00E+00 | * | 0.110 | 0.00E+00 | * |
| Cingulate | L_isthmuscingulate | 0.044 | 9.90E-02 | | 0.126 | 0.00E+00 | * | 0.098 | 6.00E-03 | * |
| Occipital | L_lateraloccipital | 0.060 | 2.60E-02 | | 0.154 | 0.00E+00 | * | -0.003 | 9.14E-01 | |
| Frontal | L_lateralorbitofrontal | 0.122 | 0.00E+00 | * | 0.245 | 0.00E+00 | * | 0.155 | 0.00E+00 | * |
| Occipital | L_lingual | 0.057 | 2.20E-02 | * | 0.111 | 2.00E-03 | * | -0.003 | 9.31E-01 | |
| Frontal | L_medialorbitofrontal | 0.068 | 1.40E-02 | * | 0.136 | 0.00E+00 | * | 0.077 | 4.30E-02 | |
| Temporal | L_middletemporal | 0.110 | 0.00E+00 | * | 0.163 | 0.00E+00 | * | 0.150 | 0.00E+00 | * |
| Temporal | L_parahippocampal | 0.034 | 2.74E-01 | | 0.071 | 4.40E-02 | | -0.100 | 4.00E-03 | * |

| | | | | | | | | | | |
|---|---|---|---|---|---|---|---|---|---|---|
| Somatosensory | L_paracentral | 0.031 | 2.93E-01 | | 0.114 | 6.00E-03 | * | -0.043 | 3.30E-01 | |
| Frontal | L_parsopercularis | 0.017 | 6.30E-01 | | 0.158 | 0.00E+00 | * | -0.060 | 2.05E-01 | |
| Frontal | L_parsorbitalis | 0.123 | 0.00E+00 | * | 0.269 | 0.00E+00 | * | 0.013 | 6.98E-01 | |
| Frontal | L_parstriangularis | 0.030 | 2.72E-01 | | 0.187 | 0.00E+00 | * | -0.047 | 1.75E-01 | |
| Occipital | L_pericalcarine | -0.011 | 6.47E-01 | | 0.069 | 1.80E-02 | * | -0.154 | 0.00E+00 | * |
| Somatosensory | L_postcentral | 0.057 | 1.42E-01 | | 0.144 | 1.00E-03 | * | 0.017 | 7.75E-01 | |
| Cingulate | L_posteriorcingulate | 0.057 | 5.30E-02 | | 0.134 | 0.00E+00 | * | 0.080 | 4.40E-02 | |
| Somatosensory | L_precentral | 0.072 | 4.10E-02 | | 0.249 | 0.00E+00 | * | 0.072 | 9.20E-02 | |
| Parietal | L_precuneus | 0.077 | 7.00E-03 | * | 0.175 | 0.00E+00 | * | 0.077 | 5.20E-02 | |
| Cingulate | L_rostralanteriorcingulate | 0.040 | 1.30E-01 | | 0.094 | 3.00E-03 | * | 0.004 | 9.31E-01 | |
| Frontal | L_rostralmiddlefrontal | 0.086 | 1.20E-02 | * | 0.200 | 0.00E+00 | * | 0.072 | 1.38E-01 | |
| Frontal | L_superiorfrontal | 0.127 | 0.00E+00 | * | 0.219 | 0.00E+00 | * | 0.138 | 0.00E+00 | * |
| Parietal | L_superiorparietal | 0.076 | 7.00E-03 | * | 0.135 | 1.00E-03 | * | 0.067 | 9.70E-02 | |
| Temporal | L_superiortemporal | 0.097 | 0.00E+00 | * | 0.174 | 0.00E+00 | * | 0.097 | 5.00E-03 | * |
| Parietal | L_supramarginal | 0.023 | 4.13E-01 | | 0.094 | 1.00E-02 | * | 0.044 | 2.23E-01 | |
| Frontal | L_frontalpole | 0.074 | 4.10E-02 | | 0.212 | 0.00E+00 | * | -0.034 | 4.68E-01 | |
| Temporal | L_temporalpole | 0.038 | 1.94E-01 | | 0.080 | 3.90E-02 | * | -0.042 | 2.80E-01 | |
| Temporal | L_transversetemporal | 0.055 | 4.30E-02 | | 0.104 | 2.00E-03 | * | 0.082 | 1.00E-02 | * |
| Insula | L_insula | 0.029 | 3.23E-01 | | 0.134 | 1.00E-03 | * | 0.079 | 3.50E-02 | |
| Temporal | R_bankssts | 0.128 | 0.00E+00 | * | 0.239 | 0.00E+00 | * | 0.031 | 3.84E-01 | |
| Cingulate | R_caudalanteriorcingulate | 0.061 | 3.00E-02 | | 0.199 | 0.00E+00 | * | 0.024 | 5.09E-01 | |
| Frontal | R_caudalmiddlefrontal | 0.022 | 4.67E-01 | | 0.084 | 2.20E-02 | * | -0.004 | 9.02E-01 | |
| Occipital | R_cuneus | -0.008 | 7.95E-01 | | -0.043 | 3.07E-01 | | -0.043 | 2.84E-01 | |
| Temporal | R_entorhinal | 0.026 | 4.26E-01 | | 0.110 | 1.60E-02 | * | -0.029 | 4.60E-01 | |
| Temporal | R_fusiform | 0.080 | 1.00E-03 | * | 0.130 | 0.00E+00 | * | 0.030 | 4.15E-01 | |
| Parietal | R_inferiorparietal | 0.062 | 1.60E-02 | * | 0.190 | 0.00E+00 | * | 0.016 | 6.46E-01 | |
| Temporal | R_inferiortemporal | 0.083 | 7.00E-03 | * | 0.144 | 0.00E+00 | * | 0.039 | 2.65E-01 | |
| Cingulate | R_isthmuscingulate | 0.080 | 5.00E-03 | * | 0.179 | 0.00E+00 | * | 0.083 | 1.90E-02 | |
| Occipital | R_lateraloccipital | 0.077 | 8.00E-03 | * | 0.181 | 0.00E+00 | * | 0.040 | 2.61E-01 | |
| Frontal | R_lateralorbitofrontal | 0.131 | 1.00E-03 | * | 0.275 | 0.00E+00 | * | 0.087 | 3.40E-02 | |
| Occipital | R_lingual | 0.060 | 2.80E-02 | | 0.084 | 6.00E-03 | * | -0.004 | 9.15E-01 | |
| Frontal | R_medialorbitofrontal | 0.115 | 0.00E+00 | * | 0.216 | 0.00E+00 | * | 0.137 | 0.00E+00 | * |
| Temporal | R_middletemporal | 0.087 | 1.00E-03 | * | 0.125 | 0.00E+00 | * | 0.128 | 2.00E-03 | * |

| | | | | | | | | | | |
|---|---|---|---|---|---|---|---|---|---|---|
| Temporal | R_parahippocampal | 0.069 | 9.00E-03 | * | 0.125 | 0.00E+00 | * | -0.002 | 9.53E-01 | |
| Somatosensory | R_paracentral | 0.006 | 8.42E-01 | | 0.050 | 1.38E-01 | | -0.047 | 2.31E-01 | |
| Frontal | R_parsopercularis | 0.103 | 1.00E-03 | * | 0.252 | 0.00E+00 | * | -0.024 | 5.16E-01 | |
| Frontal | R_parsorbitalis | 0.122 | 0.00E+00 | * | 0.228 | 0.00E+00 | * | 0.007 | 8.31E-01 | |
| Frontal | R_parstriangularis | 0.099 | 0.00E+00 | * | 0.162 | 0.00E+00 | * | -0.041 | 3.02E-01 | |
| Occipital | R_pericalcarine | 0.014 | 5.68E-01 | | 0.077 | 1.60E-02 | * | -0.106 | 0.00E+00 | * |
| Somatosensory | R_postcentral | 0.079 | 8.00E-03 | * | 0.136 | 1.00E-03 | * | 0.061 | 1.02E-01 | |
| Cingulate | R_posteriorcingulate | 0.123 | 0.00E+00 | * | 0.276 | 0.00E+00 | * | 0.115 | 2.00E-03 | * |
| Somatosensory | R_precentral | 0.155 | 0.00E+00 | * | 0.220 | 0.00E+00 | * | 0.137 | 2.00E-03 | * |
| Parietal | R_precuneus | 0.140 | 0.00E+00 | * | 0.259 | 0.00E+00 | * | 0.142 | 0.00E+00 | * |
| Cingulate | R_rostralanteriorcingulate | 0.125 | 0.00E+00 | * | 0.228 | 0.00E+00 | * | 0.068 | 5.90E-02 | |
| Frontal | R_rostralmiddlefrontal | 0.087 | 1.30E-02 | * | 0.191 | 0.00E+00 | * | 0.044 | 3.36E-01 | |
| Frontal | R_superiorfrontal | 0.122 | 0.00E+00 | * | 0.155 | 0.00E+00 | * | 0.101 | 8.00E-03 | * |
| Parietal | R_superiorparietal | 0.116 | 0.00E+00 | * | 0.170 | 0.00E+00 | * | 0.131 | 0.00E+00 | * |
| Temporal | R_superiortemporal | 0.178 | 0.00E+00 | * | 0.301 | 0.00E+00 | * | 0.153 | 0.00E+00 | * |
| Parietal | R_supramarginal | 0.126 | 0.00E+00 | * | 0.240 | 0.00E+00 | * | 0.077 | 2.60E-02 | |
| Frontal | R_frontalpole | 0.069 | 1.40E-02 | * | 0.140 | 0.00E+00 | * | 0.044 | 2.60E-01 | |
| Temporal | R_temporalpole | 0.022 | 4.51E-01 | | 0.105 | 6.00E-03 | * | -0.053 | 1.79E-01 | |
| Temporal | R_transversetemporal | 0.070 | 1.00E-02 | * | 0.180 | 0.00E+00 | * | 0.031 | 3.92E-01 | |
| Insula | R_insula | 0.062 | 3.70E-02 | | 0.159 | 0.00E+00 | * | 0.083 | 3.50E-02 | |

**Supplementary Table S5. Proportion of patients with greater deviation in regional gray matter volume measures**

| Classification | Region | All patients group | | | First-episode drug-naive group | | | Chronic medicated group | | |
|---|---|---|---|---|---|---|---|---|---|---|
| | | PD- | PD+ | (PD-)&(PD+) | PD- | PD+ | (PD-)&(PD+) | PD- | PD+ | (PD-)&(PD+) |
| Thalamus | L_thalamus | 26.1% | 11.8% | 37.8% | 22.8% | 17.8% | 40.6% | 30.4% | 8.0% | 38.4% |
| Basal Ganglia | L_caudate | 15.6% | 18.0% | 33.6% | 16.5% | 19.7% | 36.2% | 16.8% | 18.3% | 35.2% |
| Basal Ganglia | L_putamen | 14.2% | 16.6% | 30.8% | 20.7% | 18.6% | 39.3% | 14.1% | 14.6% | 28.6% |
| Basal Ganglia | L_pallidum | 11.2% | 17.0% | 28.1% | 11.1% | 13.0% | 24.1% | 10.1% | 16.6% | 26.6% |
| Hippcampus | L_hippocampus | 21.9% | 7.3% | 29.2% | 22.0% | 10.0% | 32.0% | 29.6% | 6.3% | 35.9% |
| Amygdala | L_amygdala | 19.5% | 9.8% | 29.3% | 20.7% | 7.7% | 28.5% | 22.1% | 8.0% | 30.2% |
| Basal Ganglia | L_accumbens | 21.6% | 12.7% | 34.3% | 21.1% | 12.8% | 33.9% | 23.6% | 9.3% | 32.9% |
| Thalamus | R_thalamus | 27.0% | 12.3% | 39.3% | 27.6% | 16.3% | 43.9% | 31.2% | 7.5% | 38.7% |
| Basal Ganglia | R_caudate | 14.9% | 18.8% | 33.6% | 16.1% | 18.8% | 34.9% | 16.3% | 18.3% | 34.7% |
| Basal Ganglia | R_putamen | 12.9% | 16.5% | 29.5% | 18.6% | 17.4% | 36.0% | 13.8% | 13.6% | 27.4% |
| Basal Ganglia | R_pallidum | 11.9% | 19.3% | 31.3% | 12.8% | 18.2% | 31.0% | 11.6% | 20.9% | 32.4% |
| Hippcampus | R_hippocampus | 23.0% | 9.5% | 32.6% | 20.1% | 10.9% | 31.0% | 32.2% | 7.0% | 39.2% |
| Amygdala | R_amygdala | 19.1% | 11.6% | 30.7% | 19.0% | 11.9% | 31.0% | 27.1% | 9.3% | 36.4% |
| Basal Ganglia | R_accumbens | 19.6% | 10.5% | 30.1% | 20.3% | 10.3% | 30.5% | 24.1% | 8.0% | 32.2% |
| Temporal | L_bankssts | 19.3% | 12.8% | 32.1% | 19.7% | 17.8% | 37.4% | 21.6% | 11.8% | 33.4% |
| Cingulate | L_caudalanteriorcingulate | 16.7% | 13.1% | 29.8% | 16.9% | 17.2% | 34.1% | 15.3% | 10.6% | 25.9% |
| Frontal | L_caudalmiddlefrontal | 20.7% | 12.0% | 32.7% | 20.5% | 16.9% | 37.4% | 23.4% | 11.1% | 34.4% |
| Occipital | L_cuneus | 17.1% | 12.1% | 29.1% | 16.3% | 12.8% | 29.1% | 15.3% | 8.3% | 23.6% |
| Temporal | L_entorhinal | 14.3% | 11.7% | 25.9% | 15.1% | 13.4% | 28.5% | 13.8% | 7.8% | 21.6% |
| Temporal | L_fusiform | 24.6% | 12.6% | 37.2% | 24.3% | 15.5% | 39.7% | 25.1% | 11.1% | 36.2% |
| Parietal | L_inferiorparietal | 21.0% | 12.3% | 33.3% | 21.3% | 18.2% | 39.5% | 21.4% | 8.0% | 29.4% |
| Temporal | L_inferiortemporal | 22.5% | 12.2% | 34.7% | 20.9% | 14.9% | 35.8% | 26.1% | 11.1% | 37.2% |
| Cingulate | L_isthmuscingulate | 15.2% | 11.8% | 27.0% | 18.6% | 15.1% | 33.7% | 16.6% | 11.8% | 28.4% |
| Occipital | L_lateraloccipital | 22.2% | 10.8% | 32.9% | 21.8% | 13.2% | 34.9% | 22.1% | 7.0% | 29.1% |
| Frontal | L_lateralorbitofrontal | 22.1% | 11.9% | 34.0% | 22.2% | 16.5% | 38.7% | 27.4% | 10.1% | 37.4% |
| Occipital | L_lingual | 19.9% | 11.0% | 31.0% | 18.4% | 14.0% | 32.4% | 20.9% | 7.5% | 28.4% |
| Frontal | L_medialorbitofrontal | 18.7% | 13.4% | 32.1% | 20.1% | 16.5% | 36.6% | 20.4% | 11.3% | 31.7% |
| Temporal | L_middletemporal | 23.1% | 12.9% | 36.0% | 24.3% | 18.6% | 42.9% | 25.6% | 11.3% | 36.9% |
| Temporal | L_parahippocampal | 8.4% | 6.3% | 14.7% | 8.2% | 8.6% | 16.7% | 7.5% | 3.5% | 11.1% |

| | | | | | | | | | | |
|---|---|---|---|---|---|---|---|---|---|---|
| Somatosensory | L_paracentral | 19.0% | 12.3% | 31.3% | 18.0% | 14.4% | 32.4% | 18.8% | 10.1% | 28.9% |
| Frontal | L_parsopercularis | 17.9% | 10.8% | 28.7% | 21.3% | 15.7% | 37.0% | 15.8% | 7.8% | 23.6% |
| Frontal | L_parsorbitalis | 20.6% | 13.7% | 34.3% | 22.0% | 20.5% | 42.5% | 22.6% | 8.8% | 31.4% |
| Frontal | L_parstriangularis | 19.6% | 12.9% | 32.5% | 22.0% | 21.1% | 43.1% | 17.3% | 9.5% | 26.9% |
| Occipital | L_pericalcarine | 16.0% | 14.7% | 30.7% | 15.1% | 17.4% | 32.4% | 12.8% | 11.1% | 23.9% |
| Somatosensory | L_postcentral | 22.7% | 9.8% | 32.5% | 22.8% | 13.4% | 36.2% | 24.1% | 8.5% | 32.7% |
| Cingulate | L_posteriorcingulate | 20.8% | 12.1% | 32.9% | 19.9% | 16.1% | 36.0% | 23.1% | 13.1% | 36.2% |
| Somatosensory | L_precentral | 21.8% | 9.2% | 31.0% | 22.0% | 14.0% | 36.0% | 24.6% | 8.0% | 32.7% |
| Parietal | L_precuneus | 19.5% | 11.8% | 31.3% | 20.3% | 15.9% | 36.2% | 21.1% | 9.8% | 30.9% |
| Cingulate | L_rostralanteriorcingulate | 17.0% | 11.8% | 28.8% | 18.4% | 14.9% | 33.3% | 14.3% | 9.8% | 24.1% |
| Frontal | L_rostralmiddlefrontal | 19.1% | 10.9% | 30.0% | 18.2% | 16.7% | 34.9% | 22.4% | 8.0% | 30.4% |
| Frontal | L_superiorfrontal | 24.6% | 10.8% | 35.4% | 20.5% | 15.3% | 35.8% | 29.1% | 7.0% | 36.2% |
| Parietal | L_superiorparietal | 17.7% | 12.4% | 30.1% | 17.8% | 15.7% | 33.5% | 18.3% | 9.5% | 27.9% |
| Temporal | L_superiortemporal | 24.0% | 10.4% | 34.4% | 22.2% | 14.4% | 36.6% | 26.1% | 8.3% | 34.4% |
| Parietal | L_supramarginal | 21.0% | 9.5% | 30.5% | 19.9% | 12.3% | 32.2% | 23.4% | 7.8% | 31.2% |
| Frontal | L_frontalpole | 17.2% | 13.4% | 30.6% | 17.8% | 17.4% | 35.1% | 19.3% | 8.0% | 27.4% |
| Temporal | L_temporalpole | 14.5% | 13.2% | 27.7% | 13.6% | 17.8% | 31.4% | 14.8% | 9.3% | 24.1% |
| Temporal | L_transversetemporal | 21.7% | 12.8% | 34.5% | 22.0% | 15.5% | 37.4% | 23.6% | 12.6% | 36.2% |
| Insula | L_insula | 14.9% | 5.8% | 20.7% | 16.3% | 8.6% | 24.9% | 16.1% | 5.5% | 21.6% |
| Temporal | R_bankssts | 20.8% | 12.8% | 33.6% | 19.9% | 18.8% | 38.7% | 19.8% | 10.3% | 30.2% |
| Cingulate | R_caudalanteriorcingulate | 18.5% | 14.2% | 32.7% | 22.6% | 17.6% | 40.2% | 20.6% | 11.6% | 32.2% |
| Frontal | R_caudalmiddlefrontal | 18.4% | 10.6% | 29.0% | 17.2% | 16.5% | 33.7% | 17.8% | 8.5% | 26.4% |
| Occipital | R_cuneus | 16.7% | 11.0% | 27.7% | 14.9% | 11.5% | 26.4% | 16.1% | 10.1% | 26.1% |
| Temporal | R_entorhinal | 14.4% | 13.3% | 27.7% | 15.7% | 14.2% | 29.9% | 15.6% | 11.8% | 27.4% |
| Temporal | R_fusiform | 22.9% | 11.4% | 34.3% | 21.3% | 13.6% | 34.9% | 23.9% | 8.0% | 31.9% |
| Parietal | R_inferiorparietal | 22.2% | 10.7% | 32.9% | 22.4% | 16.9% | 39.3% | 23.4% | 7.5% | 30.9% |
| Temporal | R_inferiortemporal | 22.2% | 11.3% | 33.5% | 20.5% | 16.3% | 36.8% | 24.4% | 7.3% | 31.7% |
| Cingulate | R_isthmuscingulate | 19.1% | 11.2% | 30.4% | 20.7% | 14.9% | 35.6% | 20.9% | 10.3% | 31.2% |
| Occipital | R_lateraloccipital | 21.7% | 12.2% | 33.9% | 24.5% | 14.9% | 39.3% | 23.9% | 8.5% | 32.4% |
| Frontal | R_lateralorbitofrontal | 21.0% | 13.1% | 34.1% | 20.9% | 19.9% | 40.8% | 23.9% | 8.5% | 32.4% |
| Occipital | R_lingual | 19.3% | 11.8% | 31.1% | 19.2% | 12.8% | 32.0% | 19.3% | 8.8% | 28.1% |
| Frontal | R_medialorbitofrontal | 19.4% | 9.5% | 28.9% | 19.0% | 13.2% | 32.2% | 22.4% | 6.8% | 29.1% |
| Temporal | R_middletemporal | 22.1% | 11.3% | 33.4% | 20.5% | 16.3% | 36.8% | 26.6% | 10.1% | 36.7% |

| | | | | | | | | | | |
|---|---|---|---|---|---|---|---|---|---|---|
| Temporal | R_parahippocampal | 20.5% | 12.2% | 32.7% | 20.9% | 13.2% | 34.1% | 21.1% | 8.8% | 29.9% |
| Somatosensory | R_paracentral | 20.1% | 10.7% | 30.7% | 18.4% | 13.2% | 31.6% | 19.8% | 7.0% | 26.9% |
| Frontal | R_parsopercularis | 19.4% | 11.2% | 30.6% | 19.9% | 15.5% | 35.4% | 19.8% | 7.0% | 26.9% |
| Frontal | R_parsorbitalis | 21.3% | 14.7% | 36.0% | 21.8% | 20.5% | 42.3% | 19.8% | 8.0% | 27.9% |
| Frontal | R_parstriangularis | 18.9% | 12.9% | 31.8% | 16.9% | 16.1% | 33.1% | 17.8% | 7.8% | 25.6% |
| Occipital | R_pericalcarine | 15.2% | 15.7% | 31.0% | 17.4% | 17.2% | 34.5% | 10.8% | 14.8% | 25.6% |
| Somatosensory | R_postcentral | 20.6% | 10.8% | 31.4% | 19.0% | 13.0% | 32.0% | 21.4% | 8.8% | 30.2% |
| Cingulate | R_posteriorcingulate | 23.4% | 11.7% | 35.2% | 25.9% | 16.1% | 42.1% | 23.4% | 11.1% | 34.4% |
| Somatosensory | R_precentral | 17.6% | 10.7% | 28.3% | 17.8% | 13.4% | 31.2% | 19.3% | 10.3% | 29.6% |
| Parietal | R_precuneus | 21.4% | 12.9% | 34.4% | 21.5% | 16.5% | 38.1% | 21.6% | 10.8% | 32.4% |
| Cingulate | R_rostralanteriorcingulate | 20.8% | 12.9% | 33.8% | 22.4% | 16.9% | 39.3% | 21.9% | 10.6% | 32.4% |
| Frontal | R_rostralmiddlefrontal | 16.4% | 10.4% | 26.7% | 16.7% | 15.5% | 32.2% | 19.3% | 8.3% | 27.6% |
| Frontal | R_superiorfrontal | 23.5% | 11.6% | 35.2% | 20.3% | 14.4% | 34.7% | 25.9% | 8.3% | 34.2% |
| Parietal | R_superiorparietal | 18.4% | 10.9% | 29.3% | 16.7% | 14.4% | 31.2% | 21.4% | 9.5% | 30.9% |
| Temporal | R_superiortemporal | 24.0% | 11.6% | 35.5% | 22.6% | 17.6% | 40.2% | 28.6% | 10.1% | 38.7% |
| Parietal | R_supramarginal | 19.4% | 11.8% | 31.2% | 19.5% | 15.9% | 35.4% | 20.4% | 9.8% | 30.2% |
| Frontal | R_frontalpole | 19.3% | 13.2% | 32.5% | 19.9% | 15.3% | 35.1% | 22.1% | 9.8% | 31.9% |
| Temporal | R_temporalpole | 13.8% | 14.4% | 28.2% | 14.9% | 17.6% | 32.4% | 14.6% | 10.8% | 25.4% |
| Temporal | R_transversetemporal | 22.3% | 11.6% | 33.8% | 19.7% | 16.9% | 36.6% | 25.9% | 9.0% | 34.9% |
| Insula | R_insula | 13.3% | 5.7% | 19.1% | 14.6% | 7.9% | 22.6% | 15.6% | 4.8% | 20.4% |

**Supplementary Table S6. Spatial correlation results**

| Brain map 1 | Brain map 2 | Spearman correlation test | | Spatial permutation test |
|---|---|---|---|---|
| | | r | p | pspin |
| Effect size of discovery sample | Effect size of validation sample | 0.678 | 2.72E-12 | <0.0001 |
| lnCVR | Effect size | 0.367 | 6.86E-04 | 0.0003 |
| lnCVR‖ | Effect size‖ | 0.489 | 7.42E-06 | <0.0001 |
| lnCVR | Proportion of patients in smaller volume | 0.478 | 5.61E-06 | <0.0001 |
| lnCVR | Proportion of patients in larger volume | 0.025 | 8.24E-01 | 0.5294 |

‖means that test is conducted only within regions with reduction in mean volume in patients.

**Supplementary Table S7. Correlation between brain variability map in schizophrenia and neurotransmitter map**

| Brain variability map | Neurotransmitter receptors and transporter density maps | Spearman correlation r value | Spearman correlation p value | Spin test p value |
|---|---|---|---|---|
| lnCVR | MOR | 0.3311 | 0.0024 | 0.0355 |
| PD- | 5HT2a | 0.2986 | 0.0064 | 0.0057 |
| PD- | DAT | -0.2421 | 0.0284 | 0.0219 |
| PD+ | 5HT1a | -0.2962 | 0.0069 | 0.0356 |
| PD+ | D1 | 0.3372 | 0.0019 | 0.0042 |

**Supplementary Table S8. Network diffusion model performances on estimating cross-sectional gray matter differences in schizophrenia.**

| | NDM_MC | | | NDM_SC | | | NDM_FC | | |
|---|---|---|---|---|---|---|---|---|---|
| | Model Performance(r, p) | | Optimal seed | Model Performance(r, p) | | Optimal seed | Model Performance(r, p) | | Optimal seed |
| Discovery sample (n=1,792, both early & chronic) | 0.789 | 1.00E-18 | Hippocampus | 0.547 | 1.08E-07 | Transversetemporal | 0.669 | 6.16E-12 | Hippocampus |
| Validation sample (n=4,474, predominant chronic) | 0.707 | 1.09E-13 | Parsopercularis | 0.698 | 3.19E-13 | Middletemporal | 0.723 | 1.72E-14 | Hippocampus |
| First-episode subsample (n=709) | 0.721 | 2.09E-14 | Parahippocampal | 0.487 | 4.77E-06 | Thalamus | 0.504 | 1.96E-06 | Hippocampus |
| Drug-naive subsample (n=481) | 0.432 | 6.01E-05 | Parahippocampal | 0.432 | 6.01E-05 | Thalamus | 0.431 | 6.23E-05 | Hippocampus |

* these p values are still significant through spatial permutation test and multiple comparisons correction by FDR.

**Supplementary Table S9. Model performance with each brain region as the seed of network diffusion model**

| Structure | Discovery sample (n=1,799) | | | Validation sample (n=4,474) | | | FES subsample (n=709) | | | Drug-naive subsample (n=481) | | |
|---|---|---|---|---|---|---|---|---|---|---|---|---|
| | NDM_MC | NDM_SC | NDM_FC | NDM_MC | NDM_SC | NDM_FC | NDM_MC | NDM_SC | NDM_FC | NDM_MC | NDM_SC | NDM_FC |
| Bilaterial Thalamus | 0.706 | 0.294 | 0.527 | 0.438 | 0.217 | 0.459 | 0.506 | **0.487** | **0.380** | 0.474 | **0.432** | **0.320** |
| Bilaterial Caudate | 0.368 | 0.276 | 0.378 | 0.209 | 0.018 | 0.361 | 0.446 | -0.090 | 0.083 | 0.381 | -0.122 | 0.045 |
| Bilaterial Putamen | 0.360 | 0.285 | 0.389 | 0.162 | 0.085 | 0.283 | 0.383 | 0.055 | 0.052 | 0.368 | 0.017 | 0.044 |
| Bilaterial Pallidum | 0.363 | 0.282 | -0.001 | 0.150 | 0.107 | -0.083 | 0.370 | 0.153 | 0.064 | 0.338 | 0.110 | 0.057 |
| Bilaterial Hippocampus | **0.789** | 0.340 | **0.669** | 0.593 | 0.183 | **0.723** | **0.694** | 0.225 | **0.504** | **0.623** | 0.210 | **0.431** |
| Bilaterial Amygdala | 0.719 | 0.339 | 0.153 | 0.526 | 0.267 | 0.038 | **0.641** | 0.257 | 0.108 | **0.531** | 0.225 | 0.073 |
| Bilaterial Accumbens | 0.372 | 0.185 | -0.113 | 0.237 | -0.014 | -0.099 | 0.380 | 0.075 | 0.095 | 0.369 | 0.060 | 0.104 |
| Bilaterial bankssts | 0.700 | **0.367** | 0.471 | **0.674** | **0.551** | 0.568 | 0.577 | 0.241 | 0.070 | 0.456 | 0.171 | 0.003 |
| Bilaterial caudalanteriorcingulate | 0.570 | -0.068 | 0.460 | 0.339 | -0.165 | 0.354 | 0.457 | 0.045 | 0.119 | 0.451 | 0.080 | 0.070 |
| Bilaterial caudalmiddlefrontal | 0.640 | 0.226 | 0.469 | 0.582 | 0.325 | 0.555 | 0.439 | 0.053 | 0.104 | 0.384 | -0.016 | 0.019 |
| Bilaterial cuneus | 0.491 | 0.136 | 0.455 | 0.374 | 0.217 | 0.338 | 0.470 | **0.308** | 0.231 | 0.396 | **0.291** | 0.204 |
| Bilaterial entorhinal | -0.026 | 0.008 | 0.341 | 0.505 | 0.209 | 0.250 | 0.423 | 0.227 | 0.199 | 0.392 | 0.216 | 0.158 |
| Bilaterial fusiform | **0.765** | 0.322 | **0.545** | 0.618 | 0.287 | 0.538 | 0.573 | 0.261 | **0.365** | 0.524 | 0.240 | **0.318** |
| Bilaterial inferiorparietal | 0.727 | 0.325 | 0.493 | 0.616 | **0.604** | 0.556 | 0.576 | 0.261 | 0.203 | 0.466 | 0.185 | 0.135 |
| Bilaterial inferiortemporal | **0.749** | **0.341** | 0.523 | 0.609 | **0.497** | 0.597 | 0.606 | **0.315** | **0.367** | 0.455 | 0.276 | **0.283** |
| Bilaterial isthmuscingulate | 0.614 | 0.290 | 0.410 | 0.523 | 0.020 | 0.314 | **0.622** | 0.245 | 0.147 | 0.526 | **0.286** | 0.119 |
| Bilaterial lateraloccipital | 0.586 | 0.320 | **0.546** | 0.558 | 0.321 | 0.455 | 0.489 | **0.304** | 0.319 | 0.460 | 0.269 | 0.271 |
| Bilaterial lateralorbitofrontal | 0.705 | 0.256 | 0.480 | 0.610 | 0.103 | **0.629** | 0.601 | -0.010 | 0.059 | 0.126 | -0.039 | -0.006 |
| Bilaterial lingual | 0.476 | 0.238 | 0.510 | 0.358 | 0.225 | 0.475 | 0.465 | 0.274 | 0.300 | 0.450 | 0.253 | 0.253 |
| Bilaterial medialorbitofrontal | 0.696 | 0.081 | 0.283 | 0.502 | -0.116 | 0.329 | 0.506 | -0.084 | 0.002 | 0.343 | -0.069 | -0.041 |
| Bilaterial middletemporal | 0.734 | 0.306 | 0.501 | 0.595 | **0.698** | 0.532 | 0.606 | 0.303 | 0.076 | 0.201 | 0.245 | 0.006 |
| Bilaterial parahippocampal | 0.504 | 0.231 | 0.480 | 0.571 | 0.191 | 0.329 | **0.721** | 0.241 | 0.261 | **0.675** | 0.246 | 0.210 |
| Bilaterial paracentral | 0.687 | **0.343** | 0.502 | 0.457 | -0.021 | 0.420 | 0.486 | 0.076 | 0.217 | 0.440 | 0.052 | 0.176 |
| Bilaterial parsopercularis | 0.687 | 0.150 | 0.477 | **0.707** | 0.346 | 0.559 | 0.401 | -0.031 | 0.039 | 0.212 | -0.103 | -0.011 |
| Bilaterial parsorbitalis | 0.661 | 0.112 | 0.385 | **0.621** | 0.136 | **0.641** | 0.471 | -0.104 | 0.017 | 0.439 | -0.147 | -0.061 |
| Bilaterial parstriangularis | 0.646 | 0.117 | 0.444 | **0.668** | 0.187 | **0.690** | 0.397 | -0.062 | 0.033 | 0.151 | -0.114 | -0.051 |
| Bilaterial pericalcarine | 0.484 | 0.081 | 0.429 | 0.370 | 0.216 | 0.320 | 0.448 | **0.305** | 0.203 | 0.397 | **0.288** | 0.182 |
| Bilaterial postcentral | 0.726 | **0.368** | 0.530 | 0.549 | 0.224 | 0.435 | **0.630** | 0.086 | **0.337** | **0.551** | 0.031 | **0.284** |
| Bilaterial posteriorcingulate | 0.676 | 0.279 | 0.510 | 0.464 | -0.130 | 0.485 | 0.435 | 0.084 | 0.243 | 0.518 | 0.104 | 0.172 |

| | | | | | | | | | | | | |
|---|---|---|---|---|---|---|---|---|---|---|---|---|
| Bilaterial precentral | 0.717 | 0.330 | **0.534** | 0.580 | 0.197 | 0.467 | 0.606 | 0.076 | 0.294 | 0.291 | 0.038 | 0.238 |
| Bilaterial precuneus | 0.684 | 0.292 | 0.501 | 0.591 | 0.125 | 0.451 | 0.580 | 0.283 | 0.312 | 0.239 | **0.285** | 0.243 |
| Bilaterial rostralanteriorcingulate | 0.585 | -0.056 | 0.387 | 0.386 | -0.211 | 0.284 | 0.495 | -0.062 | 0.038 | 0.410 | -0.028 | 0.007 |
| Bilaterial rostralmiddlefrontal | 0.674 | 0.283 | 0.479 | 0.533 | 0.233 | 0.526 | 0.488 | -0.088 | 0.084 | 0.098 | -0.142 | 0.011 |
| Bilaterial superiorfrontal | 0.726 | 0.303 | 0.497 | 0.581 | -0.096 | 0.583 | 0.515 | -0.100 | 0.134 | -0.026 | -0.084 | 0.072 |
| Bilaterial superiorparietal | 0.672 | 0.289 | 0.486 | 0.537 | 0.246 | 0.398 | 0.559 | 0.278 | 0.218 | 0.493 | 0.243 | 0.177 |
| Bilaterial superiortemporal | **0.755** | 0.330 | 0.513 | **0.635** | 0.449 | **0.678** | 0.581 | 0.234 | 0.305 | 0.127 | 0.177 | 0.216 |
| Bilaterial supramarginal | **0.749** | 0.324 | 0.508 | 0.619 | **0.501** | 0.492 | 0.596 | 0.141 | 0.090 | **0.539** | 0.065 | 0.046 |
| Bilaterial frontalpole | 0.623 | -0.071 | 0.248 | 0.433 | -0.054 | 0.305 | 0.471 | -0.091 | 0.034 | 0.263 | -0.088 | -0.003 |
| Bilaterial temporalpole | -0.152 | 0.060 | 0.315 | 0.484 | 0.395 | 0.314 | 0.337 | 0.285 | 0.145 | 0.321 | 0.249 | 0.098 |
| Bilaterial transversetemporal | 0.740 | **0.547** | 0.503 | 0.578 | 0.439 | 0.459 | 0.591 | 0.205 | 0.242 | 0.413 | 0.130 | 0.185 |
| Bilaterial insula | 0.666 | 0.326 | **0.564** | 0.591 | 0.465 | 0.464 | 0.590 | -0.010 | 0.171 | 0.034 | -0.085 | 0.143 |

**Supplementary Table S10. Network diffusion model (NDM)-based 'spatial' phenotypes in schizophrenia population**

| (a) NDM subgroup | Model Performance (r) | N | Age(years) | Female N (%) | PANSS N | PANSS_Pos | PANSS_Neg | PANSS_Gen | PANSS_Tot |
|---|---|---|---|---|---|---|---|---|---|
| $SZ_{nonNDM}$ | 0.42 | 500(27.9%) | 29.1(11.2) | 182(36.4%) | 314 | 19.4(7.0)* | 17.4(7.4) | 36.8(10.4)* | 73.6(20.4)* |
| $SZ_{NDM}$ | 0.48 | 1292(72.1%) | 30.2(12.1) | 578(44.7%) | 895 | 18.5(6.8)* | 17.2(7.7) | 34.9(10.4)* | 70.6(20.7)* |
| (b) Spatial phenotype | Model Performance (r) | N | | | | | | | |
| Temporal Cortex Phenotype | 0.46 | 434(24.2%) | 31.1(12.4) | 191(44.0%) | 301 | 18.3(6.8) | 17.0(7.7) | 34.1(10.3)* | 69.3(20.1)* |
| Frontal Cortex Phenotype | 0.50 | 244(13.6%) | 28.3(11.2) | 112(45.9%) | 169 | 19.0(6.6) | 17.5(7.0) | 35.2(10.1) | 71.8(19.8) |
| Cingulate-Insula Phenotype | 0.49 | 143(8.0%) | 30.5(12.7) | 59(41.3%) | 87 | 19.0(6.9) | 17.9(7.4) | 36.6(9.6) | 73.5(19.3) |
| Basal Ganglia Phenotype | 0.47 | 132(7.4%) | 31.5(13.0) | 59(44.7%) | 94 | 17.4(7.3)* | 17.0(8.3) | 34.9(12.1) | 69.3(23.9) |
| Occipital Cortex Phenotype | 0.47 | 113(6.3%) | 30.9(12.4) | 58(51.3%) | 83 | 18.9(6.5) | 18.2(8.5) | 36.3(11.6) | 73.3(23.5) |
| Parietal Cortex Phenotype | 0.49 | 102(5.7%) | 28.6(11.2) | 54(52.9%) | 64 | 18.6(6.9) | 16.6(8.8) | 33.8(9.2) | 69.0(19.1) |
| Somatosensory Cortex Phenotype | 0.49 | 72(4.0%) | 29.1(12.0) | 25(34.7%) | 53 | 18.5(7.2) | 17.1(7.7) | 35.3(10.1) | 70.8(20.7) |
| Thalamus Phenotype | 0.46 | 52(2.9%) | 31.3(12.2) | 20(38.5%) | 44 | 18.1(6.1) | 16.6(7.6) | 34.4(10.0) | 69.1(20.4) |

* P<0.05 from two-tailed two-sample t test between each subgroup and all other subgroups.

**Supplementary Table S11. Regional comparisons of gray matter volume between each spatial phenotype in schizophrenia and healthy controls**

| Region | Temporal Cortex Phenotype | | | Frontal Cortex Phenotype | | | Cingulate-Insula Phenotype | | | Basal Ganglia Phenotype | | | Occipital Cortex Phenotype | | | Parietal Cortex Phenotype | | | Somatosensory Cortex Phenotype | | | Thalamus Phenotype | | | nonNDM Phenotype | | |
|---|---|---|---|---|---|---|---|---|---|---|---|---|---|---|---|---|---|---|---|---|---|---|---|---|---|---|---|
| | T | P | ES | T | P | ES | T | P | ES | T | P | ES | T | P | ES | T | P | ES | T | P | ES | T | P | ES | T | P | ES |
| L_thalamus | -5.4 | 6.1E-08 | -0.5 | -6.8 | 2.0E-11 | -0.5 | -5.6 | 1.9E-08 | -0.3 | -5.3 | 1.2E-07 | -0.5 | -2.8 | 5.5E-03 | -0.3 | -2.6 | 1.0E-02 | -0.3 | -1.0 | 3.2E-01 | -0.1 | -7.9 | 5.6E-15 | -1.1 | -1.9 | 5.7E-02 | -0.1 |
| L_caudate | -6.0 | 2.9E-09 | -0.5 | -2.7 | 7.9E-03 | -0.2 | 3.8 | 1.7E-04 | 0.2 | 0.2 | 8.4E-01 | 0.0 | 1.3 | 1.8E-01 | 0.1 | 2.9 | 3.6E-03 | 0.3 | 2.9 | 4.2E-03 | 0.3 | -1.6 | 1.1E-01 | -0.2 | 2.9 | 4.2E-03 | 0.1 |
| L_putamen | -5.4 | 8.4E-08 | -0.5 | 0.5 | 6.5E-01 | 0.0 | 2.4 | 1.5E-02 | 0.1 | -0.6 | 5.5E-01 | -0.1 | 0.3 | 7.7E-01 | 0.0 | 2.6 | 1.0E-02 | 0.3 | 1.4 | 1.7E-01 | 0.2 | -0.7 | 4.7E-01 | -0.1 | 0.9 | 3.5E-01 | 0.0 |
| L_pallidum | -3.2 | 1.6E-03 | -0.3 | 3.8 | 1.5E-04 | 0.3 | 4.7 | 2.4E-06 | 0.3 | 1.8 | 7.3E-02 | 0.2 | 2.4 | 1.5E-02 | 0.2 | 1.7 | 9.0E-02 | 0.2 | 2.3 | 2.0E-02 | 0.3 | -1.3 | 2.1E-01 | -0.2 | 1.8 | 6.9E-02 | 0.1 |
| L_hippocampus | -3.6 | 3.7E-04 | -0.3 | -6.6 | 4.3E-11 | -0.5 | -11.8 | 0.0E+00 | -0.6 | -4.4 | 1.3E-05 | -0.4 | -5.8 | 8.6E-09 | -0.6 | -2.2 | 2.5E-02 | -0.2 | -2.9 | 3.9E-03 | -0.3 | -4.1 | 3.6E-05 | -0.6 | -5.9 | 4.9E-09 | -0.3 |
| L_amygdala | -3.6 | 3.6E-04 | -0.3 | -1.9 | 5.8E-02 | -0.1 | -8.4 | 1.1E-16 | -0.5 | -2.2 | 3.1E-02 | -0.2 | -2.0 | 4.5E-02 | -0.2 | -0.9 | 3.4E-01 | -0.1 | -2.2 | 2.6E-02 | -0.3 | -2.3 | 2.4E-02 | -0.3 | -4.2 | 2.7E-05 | -0.2 |
| L_accumbens | -8.5 | 0.0E+00 | -0.8 | -4.3 | 1.7E-05 | -0.3 | -2.5 | 1.1E-02 | -0.1 | -2.6 | 1.0E-02 | -0.2 | -1.9 | 5.5E-02 | -0.2 | 0.2 | 8.2E-01 | 0.0 | 0.1 | 9.2E-01 | 0.0 | -4.5 | 9.1E-06 | -0.6 | -3.1 | 2.3E-03 | -0.2 |
| R_thalamus | -6.6 | 5.7E-11 | -0.6 | -5.5 | 4.8E-08 | -0.4 | -6.3 | 2.9E-10 | -0.3 | -5.4 | 8.0E-08 | -0.5 | -3.8 | 1.5E-04 | -0.4 | -2.7 | 7.4E-03 | -0.3 | -1.5 | 1.3E-01 | -0.2 | -7.4 | 2.3E-13 | -1.0 | -3.2 | 1.6E-03 | -0.2 |
| R_caudate | -5.8 | 9.0E-09 | -0.5 | -3.1 | 1.9E-03 | -0.2 | 4.4 | 1.2E-05 | 0.2 | -0.4 | 7.0E-01 | 0.0 | 2.4 | 1.6E-02 | 0.2 | 3.1 | 2.1E-03 | 0.3 | 2.8 | 4.9E-03 | 0.3 | -1.4 | 1.5E-01 | -0.2 | 3.5 | 4.1E-04 | 0.2 |
| R_putamen | -4.2 | 3.2E-05 | -0.4 | 0.9 | 3.6E-01 | 0.1 | 2.5 | 1.4E-02 | 0.1 | 0.1 | 9.2E-01 | 0.0 | 1.3 | 1.9E-01 | 0.1 | 2.5 | 1.3E-02 | 0.3 | 1.7 | 8.7E-02 | 0.2 | -0.8 | 4.1E-01 | -0.1 | 1.1 | 2.9E-01 | 0.1 |
| R_pallidum | -3.1 | 2.3E-03 | -0.3 | 3.5 | 5.0E-04 | 0.2 | 4.8 | 2.1E-06 | 0.3 | 1.1 | 2.5E-01 | 0.1 | 3.0 | 3.2E-03 | 0.3 | 2.9 | 4.1E-03 | 0.3 | 2.7 | 7.2E-03 | 0.3 | -2.1 | 3.7E-02 | -0.3 | 3.6 | 2.7E-04 | 0.2 |
| R_hippocampus | -4.2 | 3.4E-05 | -0.4 | -4.9 | 1.2E-06 | -0.3 | -10.8 | 0.0E+00 | -0.6 | -4.7 | 3.0E-06 | -0.4 | -4.4 | 9.6E-06 | -0.4 | -2.2 | 2.6E-02 | -0.2 | -2.5 | 1.3E-02 | -0.3 | -3.5 | 5.6E-04 | -0.5 | -5.0 | 6.4E-07 | -0.3 |
| R_amygdala | -2.8 | 5.9E-03 | -0.3 | -1.3 | 1.8E-01 | -0.1 | -5.6 | 2.9E-08 | -0.3 | -2.2 | 2.8E-02 | -0.2 | -2.3 | 2.3E-02 | -0.2 | -0.8 | 4.4E-01 | -0.1 | -1.6 | 1.1E-01 | -0.2 | -2.0 | 4.1E-02 | -0.3 | -2.0 | 4.4E-02 | -0.1 |
| R_accumbens | -9.9 | 0.0E+00 | -0.9 | -5.4 | 8.8E-08 | -0.4 | -2.7 | 6.8E-03 | -0.1 | -2.7 | 7.4E-03 | -0.2 | -1.8 | 7.8E-02 | -0.2 | -0.6 | 5.2E-01 | -0.1 | -0.3 | 7.5E-01 | 0.0 | -4.2 | 2.4E-05 | -0.6 | -3.5 | 4.1E-04 | -0.2 |
| L_bankssts | 0.2 | 8.6E-01 | 0.0 | -3.3 | 8.8E-04 | -0.2 | -4.3 | 1.6E-05 | -0.2 | -1.7 | 9.1E-02 | -0.1 | 1.1 | 2.6E-01 | 0.1 | -3.1 | 1.9E-03 | -0.3 | -0.6 | 5.3E-01 | -0.1 | 0.3 | 7.6E-01 | 0.0 | -1.4 | 1.7E-01 | -0.1 |
| L_caudalanteriorcingulate | -1.1 | 2.6E-01 | -0.1 | -3.0 | 3.0E-03 | -0.2 | 0.0 | 9.6E-01 | 0.0 | -4.4 | 9.4E-06 | -0.4 | 0.7 | 4.7E-01 | 0.1 | -0.2 | 8.2E-01 | 0.0 | -2.1 | 3.9E-02 | -0.2 | -0.2 | 8.2E-01 | 0.0 | -2.2 | 3.0E-02 | -0.1 |
| L_caudalmiddlefrontal | -3.3 | 1.1E-03 | -0.3 | -6.5 | 7.7E-11 | -0.5 | -2.0 | 4.2E-02 | -0.1 | -3.7 | 2.1E-04 | -0.3 | -1.2 | 2.3E-01 | -0.1 | -2.3 | 2.1E-02 | -0.2 | -3.0 | 3.1E-03 | -0.4 | -2.0 | 4.7E-02 | -0.3 | 0.9 | 3.5E-01 | 0.0 |
| L_cuneus | -1.2 | 2.4E-01 | -0.1 | 0.0 | 1.0E+00 | 0.0 | -0.3 | 7.6E-01 | 0.0 | 0.2 | 8.1E-01 | 0.0 | -10.0 | 0.0E+00 | -1.0 | -4.4 | 1.3E-05 | -0.4 | 1.2 | 2.2E-01 | 0.1 | -2.4 | 1.5E-02 | -0.3 | -1.1 | 2.9E-01 | -0.1 |
| L_entorhinal | 0.3 | 7.6E-01 | 0.0 | 1.9 | 6.1E-02 | 0.1 | -4.3 | 1.6E-05 | -0.2 | -0.9 | 3.9E-01 | -0.1 | -0.7 | 4.7E-01 | -0.1 | 0.1 | 9.1E-01 | 0.0 | 1.4 | 1.8E-01 | 0.2 | 0.8 | 4.1E-01 | 0.1 | -2.8 | 4.9E-03 | -0.1 |
| L_fusiform | -2.5 | 1.4E-02 | -0.2 | -5.7 | 1.8E-08 | -0.4 | -9.6 | 0.0E+00 | -0.5 | -3.0 | 2.5E-03 | -0.3 | -2.3 | 2.1E-02 | -0.2 | -1.7 | 8.7E-02 | -0.2 | -1.6 | 1.1E-01 | -0.2 | -1.0 | 3.1E-01 | -0.1 | -2.1 | 3.3E-02 | -0.1 |
| L_inferiorparietal | -1.3 | 1.8E-01 | -0.1 | -6.0 | 2.7E-09 | -0.4 | -3.9 | 8.8E-05 | -0.2 | -4.3 | 1.6E-05 | -0.4 | -2.0 | 4.8E-02 | -0.2 | -3.4 | 6.9E-04 | -0.3 | -0.3 | 7.3E-01 | 0.0 | -0.7 | 4.9E-01 | -0.1 | -2.7 | 7.3E-03 | -0.1 |
| L_inferiortemporal | -2.4 | 1.8E-02 | -0.2 | -6.0 | 1.8E-09 | -0.4 | -6.7 | 2.3E-11 | -0.4 | -4.3 | 1.7E-05 | -0.4 | -2.2 | 3.1E-02 | -0.2 | -1.9 | 5.8E-02 | -0.2 | -0.8 | 4.1E-01 | -0.1 | 1.7 | 8.6E-02 | 0.2 | -2.8 | 4.5E-03 | -0.1 |
| L_isthmuscingulate | 0.4 | 6.8E-01 | 0.0 | -2.2 | 3.1E-02 | -0.1 | -1.1 | 2.6E-01 | -0.1 | -4.3 | 1.7E-05 | -0.4 | -4.2 | 2.4E-05 | -0.4 | -4.7 | 3.0E-06 | -0.5 | 0.7 | 4.8E-01 | 0.1 | -1.5 | 1.3E-01 | -0.2 | -0.1 | 9.5E-01 | 0.0 |
| L_lateraloccipital | -3.0 | 2.4E-03 | -0.3 | -4.7 | 2.5E-06 | -0.3 | -3.7 | 1.8E-04 | -0.2 | -2.6 | 8.3E-03 | -0.2 | -7.0 | 3.4E-12 | -0.7 | -4.4 | 1.4E-05 | -0.4 | -3.0 | 3.1E-03 | -0.4 | -1.5 | 1.4E-01 | -0.2 | -1.9 | 6.2E-02 | -0.1 |
| L_lateralorbitofrontal | -3.4 | 7.5E-04 | -0.3 | -9.9 | 0.0E+00 | -0.7 | -3.9 | 9.7E-05 | -0.2 | -5.7 | 1.3E-08 | -0.5 | -0.8 | 4.0E-01 | -0.1 | -2.6 | 9.4E-03 | -0.3 | -3.9 | 1.2E-04 | -0.5 | -0.7 | 4.7E-01 | -0.1 | -0.8 | 4.0E-01 | 0.0 |
| L_lingual | -2.6 | 8.6E-03 | -0.2 | -0.5 | 6.3E-01 | 0.0 | -2.7 | 7.8E-03 | -0.1 | -1.0 | 3.1E-01 | -0.1 | -10.7 | 0.0E+00 | -1.0 | -3.9 | 1.1E-04 | -0.4 | -1.0 | 3.4E-01 | -0.1 | -3.2 | 1.3E-03 | -0.5 | -1.8 | 7.8E-02 | -0.1 |
| L_medialorbitofrontal | -1.1 | 2.7E-01 | -0.1 | -6.8 | 1.3E-11 | -0.5 | -2.6 | 1.0E-02 | -0.1 | -2.0 | 4.6E-02 | -0.2 | 0.9 | 3.9E-01 | 0.1 | 0.0 | 9.7E-01 | 0.0 | -2.3 | 2.5E-02 | -0.3 | -0.4 | 7.2E-01 | -0.1 | -0.5 | 6.1E-01 | 0.0 |
| L_middletemporal | -2.3 | 2.0E-02 | -0.2 | -8.2 | 5.6E-16 | -0.6 | -5.7 | 1.7E-08 | -0.3 | -4.8 | 2.2E-06 | -0.4 | -2.5 | 1.3E-02 | -0.2 | -2.6 | 8.9E-03 | -0.3 | -0.4 | 7.3E-01 | 0.0 | 0.3 | 7.9E-01 | 0.0 | -0.8 | 4.2E-01 | 0.0 |
| L_parahippocampal | 0.3 | 7.8E-01 | 0.0 | 0.0 | 1.0E+00 | 0.0 | -7.8 | 7.9E-15 | -0.4 | -1.1 | 2.7E-01 | -0.1 | -1.9 | 6.3E-02 | -0.2 | 0.3 | 7.7E-01 | 0.0 | -0.8 | 4.3E-01 | -0.1 | -1.1 | 2.6E-01 | -0.2 | -2.7 | 7.3E-03 | -0.1 |
| L_paracentral | -1.8 | 7.7E-02 | -0.2 | -5.6 | 3.1E-08 | -0.4 | -2.6 | 1.1E-02 | -0.1 | -3.2 | 1.6E-03 | -0.3 | -0.1 | 9.5E-01 | 0.0 | -3.0 | 3.2E-03 | -0.3 | -5.4 | 8.5E-08 | -0.6 | -2.2 | 2.6E-02 | -0.3 | 0.1 | 8.8E-01 | 0.0 |
| L_parsopercularis | -3.0 | 3.0E-03 | -0.3 | -7.3 | 5.0E-13 | -0.5 | -1.3 | 1.9E-01 | -0.1 | -3.8 | 1.6E-04 | -0.3 | -2.4 | 1.7E-02 | -0.2 | -1.4 | 1.7E-01 | -0.1 | -1.9 | 5.5E-02 | -0.2 | -1.0 | 3.2E-01 | -0.1 | -1.4 | 1.5E-01 | -0.1 |
| L_parsorbitalis | -2.8 | 4.7E-03 | -0.3 | -9.2 | 0.0E+00 | -0.6 | -2.8 | 5.5E-03 | -0.2 | -2.5 | 1.4E-02 | -0.2 | -2.3 | 2.2E-02 | -0.2 | -0.5 | 6.0E-01 | -0.1 | -2.3 | 2.4E-02 | -0.3 | -0.6 | 5.6E-01 | -0.1 | -0.1 | 9.2E-01 | 0.0 |
| L_parstriangularis | -2.0 | 4.5E-02 | -0.2 | -7.7 | 1.8E-14 | -0.5 | -1.9 | 6.2E-02 | -0.1 | -2.6 | 8.3E-03 | -0.2 | -1.9 | 5.7E-02 | -0.2 | -0.9 | 3.9E-01 | -0.1 | 0.4 | 6.9E-01 | 0.0 | -2.0 | 4.3E-02 | -0.3 | -1.0 | 3.2E-01 | -0.1 |
| L_pericalcarine | 0.7 | 5.1E-01 | 0.1 | 2.2 | 2.5E-02 | 0.2 | 2.4 | 1.5E-02 | 0.1 | -0.3 | 7.8E-01 | 0.0 | -9.8 | 0.0E+00 | -1.0 | -2.0 | 4.9E-02 | -0.2 | 0.8 | 4.3E-01 | 0.1 | -1.9 | 5.5E-02 | -0.3 | -0.4 | 6.7E-01 | 0.0 |
| L_postcentral | -3.7 | 2.2E-04 | -0.3 | -5.4 | 9.2E-08 | -0.4 | -6.6 | 5.7E-11 | -0.4 | -4.7 | 3.1E-06 | -0.4 | -2.2 | 2.8E-02 | -0.2 | -4.7 | 3.0E-06 | -0.5 | -5.6 | 3.2E-08 | -0.7 | -1.8 | 6.9E-02 | -0.3 | -0.8 | 4.3E-01 | 0.0 |
| L_posteriorcingulate | -2.3 | 2.1E-02 | -0.2 | -5.6 | 2.0E-08 | -0.4 | -3.2 | 1.5E-03 | -0.2 | -5.6 | 2.0E-08 | -0.5 | 0.4 | 6.8E-01 | 0.0 | -3.9 | 8.6E-05 | -0.4 | -2.7 | 8.0E-03 | -0.3 | -1.8 | 7.0E-02 | -0.3 | -1.0 | 3.0E-01 | -0.1 |
| L_precentral | -4.4 | 1.1E-05 | -0.4 | -6.7 | 3.2E-11 | -0.5 | -5.0 | 5.2E-07 | -0.3 | -4.5 | 7.8E-06 | -0.4 | -2.7 | 7.7E-03 | -0.3 | -4.7 | 2.9E-06 | -0.5 | -5.9 | 5.6E-09 | -0.7 | -3.4 | 6.5E-04 | -0.5 | -2.4 | 1.7E-02 | -0.1 |
| L_precuneus | -3.1 | 2.0E-03 | -0.3 | -4.8 | 2.1E-06 | -0.3 | -2.4 | 1.5E-02 | -0.1 | -5.6 | 3.0E-08 | -0.5 | -1.3 | 1.8E-01 | -0.1 | -6.1 | 1.2E-09 | -0.6 | -1.1 | 2.9E-01 | -0.1 | -1.0 | 3.3E-01 | -0.1 | -0.3 | 7.5E-01 | 0.0 |
| L_rostralanteriorcingulate | -2.3 | 2.3E-02 | -0.2 | -4.1 | 3.6E-05 | -0.3 | -1.7 | 8.2E-02 | -0.1 | -5.6 | 2.5E-08 | -0.5 | -0.2 | 8.2E-01 | 0.0 | -0.2 | 8.7E-01 | 0.0 | -1.3 | 1.8E-01 | -0.2 | 0.7 | 5.1E-01 | 0.1 | -2.0 | 4.1E-02 | -0.1 |
| L_rostralmiddlefrontal | -2.2 | 2.6E-02 | -0.2 | -9.8 | 0.0E+00 | -0.7 | -2.8 | 6.0E-03 | -0.1 | -5.2 | 2.3E-07 | -0.5 | -2.2 | 2.6E-02 | -0.2 | -0.5 | 5.9E-01 | -0.1 | -0.6 | 5.5E-01 | -0.1 | 0.8 | 4.5E-01 | 0.1 | -1.0 | 3.2E-01 | -0.1 |
| L_superiorfrontal | -3.1 | 2.2E-03 | -0.3 | -12.3 | 0.0E+00 | -0.8 | -5.1 | 4.4E-07 | -0.3 | -6.3 | 3.1E-10 | -0.6 | -2.9 | 4.3E-03 | -0.3 | -4.1 | 4.9E-05 | -0.4 | -2.7 | 6.7E-03 | -0.3 | -1.0 | 3.4E-01 | -0.1 | -1.0 | 3.3E-01 | -0.1 |
| L_superiorparietal | -2.5 | 1.4E-02 | -0.2 | -3.5 | 5.6E-04 | -0.2 | -1.7 | 8.0E-02 | -0.1 | -3.8 | 1.6E-04 | -0.3 | -0.3 | 7.3E-01 | 0.0 | -5.1 | 4.9E-07 | -0.5 | -1.7 | 9.4E-02 | -0.2 | -1.5 | 1.3E-01 | -0.2 | -0.9 | 3.5E-01 | 0.0 |
| L_superiortemporal | -2.6 | 9.2E-03 | -0.2 | -7.1 | 2.2E-12 | -0.5 | -7.7 | 2.5E-14 | -0.4 | -4.4 | 1.1E-05 | -0.4 | -0.7 | 4.9E-01 | -0.1 | -2.7 | 7.8E-03 | -0.3 | -3.5 | 5.1E-04 | -0.4 | -1.6 | 1.2E-01 | -0.2 | -2.5 | 1.4E-02 | -0.1 |
| L_supramarginal | -1.7 | 8.4E-02 | -0.2 | -4.6 | 4.9E-06 | -0.3 | -5.3 | 1.4E-07 | -0.3 | -4.8 | 1.4E-06 | -0.4 | -1.9 | 5.9E-02 | -0.2 | -4.6 | 4.5E-06 | -0.5 | -1.8 | 6.8E-02 | -0.2 | -2.7 | 6.2E-03 | -0.4 | -2.1 | 3.7E-02 | -0.1 |
| L_frontalpole | 1.8 | 6.8E-02 | 0.2 | -8.3 | 2.2E-16 | -0.6 | 0.5 | 6.0E-01 | 0.0 | -0.9 | 3.7E-01 | -0.1 | -1.2 | 2.3E-01 | -0.1 | 0.2 | 8.4E-01 | 0.0 | 0.3 | 7.9E-01 | 0.0 | 1.3 | 1.9E-01 | 0.2 | -2.0 | 4.4E-02 | -0.1 |

| | | | | | | | | | | | | | | | | | | | | | | | | | | | |
|---|---|---|---|---|---|---|---|---|---|---|---|---|---|---|---|---|---|---|---|---|---|---|---|---|---|---|---|
| L_temporalpole | 1.4 | 1.6E-01 | 0.1 | 1.7 | 9.2E-02 | 0.1 | -5.0 | 6.5E-07 | -0.3 | 0.4 | 7.1E-01 | 0.0 | 0.8 | 4.1E-01 | 0.1 | 3.4 | 6.1E-04 | 0.4 | 0.9 | 3.7E-01 | 0.1 | 1.1 | 2.8E-01 | 0.2 | -1.5 | 1.3E-01 | -0.1 |
| L_transversetemporal | -1.6 | 1.2E-01 | -0.1 | -2.8 | 4.6E-03 | -0.2 | -4.5 | 7.7E-06 | -0.2 | -3.2 | 1.4E-03 | -0.3 | -0.9 | 3.7E-01 | -0.1 | -3.1 | 2.1E-03 | -0.3 | -2.7 | 8.0E-03 | -0.3 | -1.6 | 1.2E-01 | -0.2 | -1.5 | 1.4E-01 | -0.1 |
| L_insula | -3.8 | 1.6E-04 | -0.3 | -7.6 | 3.8E-14 | -0.5 | -5.1 | 2.9E-07 | -0.3 | -6.9 | 6.9E-12 | -0.6 | -1.7 | 8.6E-02 | -0.2 | -2.1 | 3.8E-02 | -0.2 | -2.1 | 3.6E-02 | -0.3 | -1.9 | 5.5E-02 | -0.3 | -3.5 | 4.7E-04 | -0.2 |
| R_bankssts | -2.9 | 3.8E-03 | -0.3 | -5.0 | 5.4E-07 | -0.3 | -5.1 | 3.7E-07 | -0.3 | -2.9 | 3.5E-03 | -0.3 | -1.0 | 3.3E-01 | -0.1 | -1.8 | 7.1E-02 | -0.2 | -2.7 | 6.1E-03 | -0.3 | -0.2 | 8.7E-01 | 0.0 | -0.5 | 5.8E-01 | 0.0 |
| R_caudalanteriorcingulate | -3.0 | 3.1E-03 | -0.3 | -2.2 | 2.8E-02 | -0.2 | 0.0 | 9.9E-01 | 0.0 | -6.8 | 1.5E-11 | -0.6 | 1.0 | 3.0E-01 | 0.1 | 1.6 | 1.1E-01 | 0.2 | -1.7 | 8.3E-02 | -0.2 | 0.5 | 5.9E-01 | 0.1 | -2.2 | 3.2E-02 | -0.1 |
| R_caudalmiddlefrontal | -2.6 | 1.0E-02 | -0.2 | -6.3 | 4.7E-10 | -0.4 | -2.3 | 2.0E-02 | -0.1 | -3.6 | 2.9E-04 | -0.3 | -1.1 | 2.6E-01 | -0.1 | -1.8 | 6.8E-02 | -0.2 | -2.4 | 1.5E-02 | -0.3 | -2.3 | 2.0E-02 | -0.3 | 1.5 | 1.3E-01 | 0.1 |
| R_cuneus | -1.4 | 1.6E-01 | -0.1 | -0.8 | 4.0E-01 | -0.1 | -1.5 | 1.2E-01 | -0.1 | -0.3 | 7.8E-01 | 0.0 | -8.9 | 0.0E+00 | -0.9 | -4.5 | 7.8E-06 | -0.5 | -0.8 | 4.4E-01 | -0.1 | -1.6 | 1.1E-01 | -0.2 | -1.9 | 5.5E-02 | -0.1 |
| R_entorhinal | 1.1 | 2.9E-01 | 0.1 | 1.5 | 1.4E-01 | 0.1 | -5.7 | 1.2E-08 | -0.3 | -0.1 | 9.4E-01 | 0.0 | 0.6 | 5.8E-01 | 0.1 | 1.1 | 2.9E-01 | 0.1 | 0.9 | 3.6E-01 | 0.1 | 1.1 | 2.9E-01 | 0.2 | -2.2 | 3.1E-02 | -0.1 |
| R_fusiform | -2.8 | 5.2E-03 | -0.3 | -5.0 | 7.4E-07 | -0.3 | -9.9 | 0.0E+00 | -0.5 | -3.3 | 1.2E-03 | -0.3 | -3.0 | 2.4E-03 | -0.3 | -2.0 | 4.3E-02 | -0.2 | -3.4 | 7.3E-04 | -0.4 | -0.6 | 5.4E-01 | -0.1 | -2.7 | 6.4E-03 | -0.1 |
| R_inferiorparietal | -0.6 | 5.8E-01 | -0.1 | -6.8 | 1.3E-11 | -0.5 | -5.9 | 4.3E-09 | -0.3 | -4.7 | 3.4E-06 | -0.4 | -1.9 | 5.5E-02 | -0.2 | -4.1 | 4.3E-05 | -0.4 | -0.8 | 4.0E-01 | -0.1 | -2.0 | 4.5E-02 | -0.3 | -1.7 | 9.6E-02 | -0.1 |
| R_inferiortemporal | -2.3 | 2.3E-02 | -0.2 | -5.3 | 1.0E-07 | -0.4 | -8.5 | 0.0E+00 | -0.5 | -3.0 | 3.2E-03 | -0.3 | 0.0 | 9.7E-01 | 0.0 | -1.4 | 1.5E-01 | -0.1 | -1.2 | 2.3E-01 | -0.1 | 1.2 | 2.3E-01 | 0.2 | -3.8 | 1.7E-04 | -0.2 |
| R_isthmuscingulate | 0.4 | 6.8E-01 | 0.0 | -2.2 | 2.7E-02 | -0.2 | -2.4 | 1.4E-02 | -0.1 | -4.3 | 1.5E-05 | -0.4 | -4.7 | 2.9E-06 | -0.5 | -5.1 | 4.5E-07 | -0.5 | 0.8 | 4.1E-01 | 0.1 | -1.2 | 2.4E-01 | -0.2 | -1.9 | 6.1E-02 | -0.1 |
| R_lateraloccipital | -1.8 | 7.9E-02 | -0.2 | -3.7 | 2.2E-04 | -0.3 | -4.0 | 5.4E-05 | -0.2 | -3.2 | 1.4E-03 | -0.3 | -7.7 | 2.7E-14 | -0.7 | -4.3 | 1.5E-05 | -0.4 | -3.8 | 1.6E-04 | -0.5 | -2.0 | 4.7E-02 | -0.3 | -1.9 | 6.1E-02 | -0.1 |
| R_lateralorbitofrontal | -2.3 | 2.4E-02 | -0.2 | -9.0 | 0.0E+00 | -0.6 | -2.7 | 6.4E-03 | -0.1 | -4.8 | 1.7E-06 | -0.4 | -1.3 | 1.8E-01 | -0.1 | -1.6 | 1.1E-01 | -0.2 | -2.5 | 1.1E-02 | -0.3 | 1.3 | 1.8E-01 | 0.2 | -1.1 | 2.9E-01 | -0.1 |
| R_lingual | -1.7 | 9.6E-02 | -0.2 | -0.6 | 5.2E-01 | 0.0 | -1.6 | 1.1E-01 | -0.1 | -1.9 | 5.1E-02 | -0.2 | -11.0 | 0.0E+00 | -1.1 | -3.7 | 2.2E-04 | -0.4 | -1.2 | 2.1E-01 | -0.1 | -2.5 | 1.2E-02 | -0.4 | -1.5 | 1.3E-01 | -0.1 |
| R_medialorbitofrontal | -4.2 | 2.7E-05 | -0.4 | -9.1 | 0.0E+00 | -0.6 | -4.1 | 3.9E-05 | -0.2 | -5.1 | 3.7E-07 | -0.4 | -2.0 | 4.1E-02 | -0.2 | -1.0 | 3.0E-01 | -0.1 | -1.4 | 1.6E-01 | -0.2 | 0.7 | 4.8E-01 | 0.1 | -2.5 | 1.4E-02 | -0.1 |
| R_middletemporal | -2.0 | 5.0E-02 | -0.2 | -8.3 | 3.3E-16 | -0.6 | -6.8 | 1.4E-11 | -0.4 | -4.1 | 3.9E-05 | -0.4 | -2.5 | 1.2E-02 | -0.2 | -3.0 | 2.4E-03 | -0.3 | -0.5 | 6.4E-01 | -0.1 | -0.1 | 9.1E-01 | 0.0 | -0.8 | 4.1E-01 | 0.0 |
| R_parahippocampal | -1.4 | 1.6E-01 | -0.1 | 0.2 | 8.2E-01 | 0.0 | -7.4 | 2.5E-13 | -0.4 | -1.5 | 1.4E-01 | -0.1 | -3.3 | 8.9E-04 | -0.3 | -1.5 | 1.2E-01 | -0.2 | -1.4 | 1.6E-01 | -0.2 | -2.6 | 8.9E-03 | -0.4 | -2.9 | 3.8E-03 | -0.1 |
| R_paracentral | -3.8 | 1.6E-04 | -0.3 | -5.2 | 2.6E-07 | -0.4 | -4.1 | 4.9E-05 | -0.2 | -5.3 | 1.3E-07 | -0.5 | -0.6 | 5.2E-01 | -0.1 | -1.9 | 5.4E-02 | -0.2 | -6.4 | 2.0E-10 | -0.8 | -3.9 | 8.4E-05 | -0.6 | -0.2 | 8.2E-01 | 0.0 |
| R_parsopercularis | -2.3 | 2.4E-02 | -0.2 | -7.9 | 4.0E-15 | -0.5 | -1.7 | 8.9E-02 | -0.1 | -3.3 | 1.1E-03 | -0.3 | -2.5 | 1.4E-02 | -0.2 | -0.1 | 9.4E-01 | 0.0 | -0.8 | 4.3E-01 | -0.1 | -1.1 | 2.5E-01 | -0.2 | -1.9 | 5.6E-02 | -0.1 |
| R_parsorbitalis | -3.1 | 2.3E-03 | -0.3 | -9.3 | 0.0E+00 | -0.6 | -1.5 | 1.5E-01 | -0.1 | -1.2 | 2.4E-01 | -0.1 | -2.3 | 2.0E-02 | -0.2 | 0.6 | 5.3E-01 | 0.1 | -0.5 | 6.5E-01 | -0.1 | 2.6 | 9.0E-03 | 0.4 | -0.8 | 4.2E-01 | 0.0 |
| R_parstriangularis | -2.0 | 5.1E-02 | -0.2 | -7.8 | 1.0E-14 | -0.5 | -2.0 | 4.2E-02 | -0.1 | -2.5 | 1.3E-02 | -0.2 | -1.6 | 1.0E-01 | -0.2 | 1.0 | 3.2E-01 | 0.1 | 2.1 | 3.3E-02 | 0.3 | 0.7 | 4.8E-01 | 0.1 | -1.6 | 1.2E-01 | -0.1 |
| R_pericalcarine | 1.0 | 3.2E-01 | 0.1 | 2.3 | 2.1E-02 | 0.2 | 2.7 | 7.9E-03 | 0.1 | 0.2 | 8.3E-01 | 0.0 | -9.7 | 0.0E+00 | -1.0 | -2.1 | 4.0E-02 | -0.2 | 0.6 | 5.5E-01 | 0.1 | -1.8 | 7.7E-02 | -0.2 | -0.3 | 7.7E-01 | 0.0 |
| R_postcentral | -0.6 | 5.7E-01 | -0.1 | -4.6 | 5.1E-06 | -0.3 | -3.6 | 3.1E-04 | -0.2 | -3.9 | 8.3E-05 | -0.3 | -2.6 | 9.4E-03 | -0.3 | -3.8 | 1.3E-04 | -0.4 | -6.8 | 1.7E-11 | -0.8 | -2.6 | 8.3E-03 | -0.4 | -1.6 | 1.0E-01 | -0.1 |
| R_posteriorcingulate | -3.9 | 8.2E-05 | -0.4 | -5.5 | 4.0E-08 | -0.4 | -4.1 | 4.2E-05 | -0.2 | -8.5 | 0.0E+00 | -0.7 | -1.1 | 2.7E-01 | -0.1 | -2.2 | 2.9E-02 | -0.2 | -4.5 | 6.5E-06 | -0.5 | -1.8 | 6.8E-02 | -0.3 | -1.8 | 6.8E-02 | -0.1 |
| R_precentral | -1.5 | 1.3E-01 | -0.1 | -6.0 | 2.8E-09 | -0.4 | -4.4 | 1.1E-05 | -0.2 | -4.6 | 5.4E-06 | -0.4 | -1.9 | 6.4E-02 | -0.2 | -4.0 | 7.1E-05 | -0.4 | -6.8 | 1.1E-11 | -0.8 | -3.6 | 2.8E-04 | -0.5 | -1.0 | 3.0E-01 | -0.1 |
| R_precuneus | -1.9 | 5.5E-02 | -0.2 | -5.0 | 7.9E-07 | -0.3 | -2.9 | 4.1E-03 | -0.2 | -5.7 | 1.6E-08 | -0.5 | -2.1 | 3.3E-02 | -0.2 | -8.1 | 7.8E-16 | -0.8 | -1.7 | 8.3E-02 | -0.2 | -1.3 | 1.8E-01 | -0.2 | -0.7 | 4.9E-01 | 0.0 |
| R_rostralanteriorcingulate | -2.1 | 3.5E-02 | -0.2 | -4.8 | 1.4E-06 | -0.3 | -2.9 | 4.3E-03 | -0.2 | -7.7 | 3.0E-14 | -0.7 | 1.3 | 2.1E-01 | 0.1 | 0.7 | 4.6E-01 | 0.1 | -2.5 | 1.1E-02 | -0.3 | 0.8 | 4.1E-01 | 0.1 | -2.0 | 4.2E-02 | -0.1 |
| R_rostralmiddlefrontal | -2.5 | 1.2E-02 | -0.2 | -8.5 | 0.0E+00 | -0.6 | -1.8 | 7.4E-02 | -0.1 | -3.8 | 1.6E-04 | -0.3 | -1.6 | 1.1E-01 | -0.2 | -0.2 | 8.7E-01 | 0.0 | 1.9 | 5.5E-02 | 0.2 | 1.7 | 8.6E-02 | 0.2 | -1.5 | 1.4E-01 | -0.1 |
| R_superiorfrontal | -4.1 | 4.0E-05 | -0.4 | -11.2 | 0.0E+00 | -0.8 | -4.8 | 1.9E-06 | -0.3 | -5.1 | 3.3E-07 | -0.4 | -2.4 | 1.8E-02 | -0.2 | -2.5 | 1.3E-02 | -0.3 | -1.2 | 2.2E-01 | -0.1 | -1.6 | 1.0E-01 | -0.2 | -0.4 | 7.0E-01 | 0.0 |
| R_superiorparietal | -2.5 | 1.2E-02 | -0.2 | -4.4 | 9.1E-06 | -0.3 | -2.0 | 4.3E-02 | -0.1 | -3.8 | 1.5E-04 | -0.3 | 0.0 | 9.8E-01 | 0.0 | -6.9 | 1.0E-11 | -0.7 | -2.4 | 1.5E-02 | -0.3 | -2.3 | 2.1E-02 | -0.3 | -1.1 | 2.6E-01 | -0.1 |
| R_superiortemporal | -3.6 | 3.5E-04 | -0.3 | -8.1 | 1.2E-15 | -0.6 | -6.5 | 1.0E-10 | -0.4 | -6.3 | 3.5E-10 | -0.6 | -1.3 | 2.0E-01 | -0.1 | -2.1 | 3.5E-02 | -0.2 | -2.7 | 7.9E-03 | -0.3 | -0.4 | 6.9E-01 | -0.1 | -1.9 | 5.6E-02 | -0.1 |
| R_supramarginal | -0.8 | 4.0E-01 | -0.1 | -3.7 | 2.2E-04 | -0.3 | -4.9 | 1.3E-06 | -0.3 | -4.4 | 1.4E-05 | -0.4 | -0.6 | 5.3E-01 | -0.1 | -4.7 | 2.7E-06 | -0.5 | -4.1 | 4.7E-05 | -0.5 | -1.2 | 2.1E-01 | -0.2 | -1.9 | 6.0E-02 | -0.1 |
| R_frontalpole | 0.2 | 8.3E-01 | 0.0 | -8.3 | 1.1E-16 | -0.6 | 0.5 | 6.3E-01 | 0.0 | -1.9 | 6.4E-02 | -0.2 | -0.6 | 5.4E-01 | -0.1 | -0.4 | 7.3E-01 | 0.0 | 0.6 | 5.5E-01 | 0.1 | 0.9 | 3.9E-01 | 0.1 | -3.7 | 1.9E-04 | -0.2 |
| R_temporalpole | 3.0 | 2.5E-03 | 0.3 | 0.4 | 6.7E-01 | 0.0 | -4.4 | 1.3E-05 | -0.2 | 0.1 | 8.9E-01 | 0.0 | 2.2 | 3.1E-02 | 0.2 | 2.5 | 1.4E-02 | 0.3 | 0.8 | 4.1E-01 | 0.1 | 1.1 | 2.9E-01 | 0.2 | -1.9 | 5.7E-02 | -0.1 |
| R_transversetemporal | -2.8 | 5.2E-03 | -0.3 | -3.9 | 8.6E-05 | -0.3 | -4.8 | 1.4E-06 | -0.3 | -3.7 | 2.3E-04 | -0.3 | -1.8 | 7.6E-02 | -0.2 | -4.5 | 6.0E-06 | -0.5 | -2.8 | 4.7E-03 | -0.3 | 0.3 | 7.8E-01 | 0.0 | -4.0 | 7.7E-05 | -0.2 |
| R_insula | -3.3 | 1.2E-03 | -0.3 | -7.2 | 6.9E-13 | -0.5 | -5.1 | 3.7E-07 | -0.3 | -6.1 | 1.7E-09 | -0.5 | -2.1 | 3.8E-02 | -0.2 | -0.5 | 5.9E-01 | -0.1 | -2.6 | 1.1E-02 | -0.3 | -2.7 | 6.9E-03 | -0.4 | -3.2 | 1.3E-03 | -0.2 |

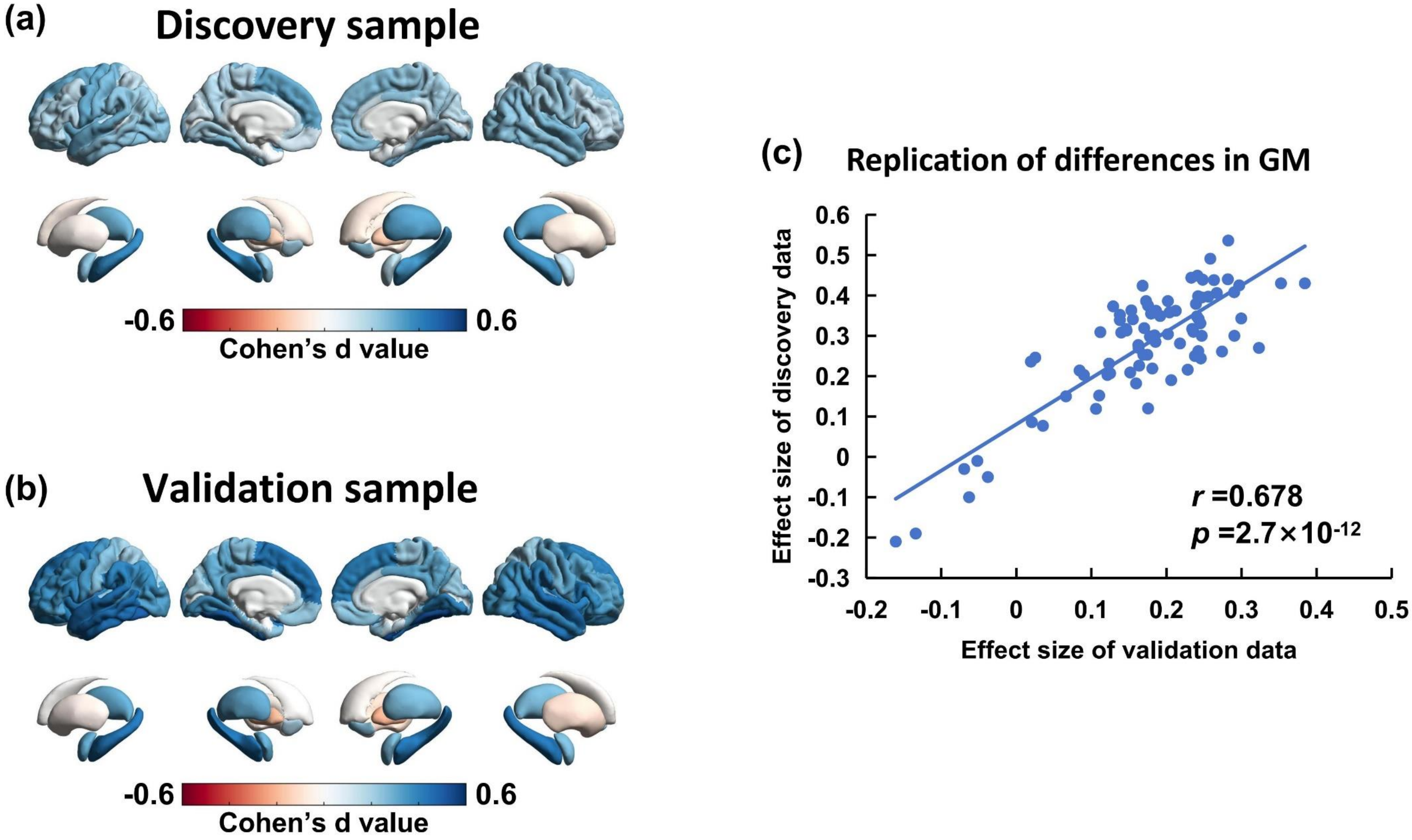


**Supplementary Figure 1.** High consistency between the case-control difference (i.e., effect size) brain map that sourced from **(a)** discovery sample and **(b)** validation sample. **(c)** It shows a highly spatial correlation ($r$=0.678, $p$=2.7×$10^{-12}$) between discovery data and validation data.

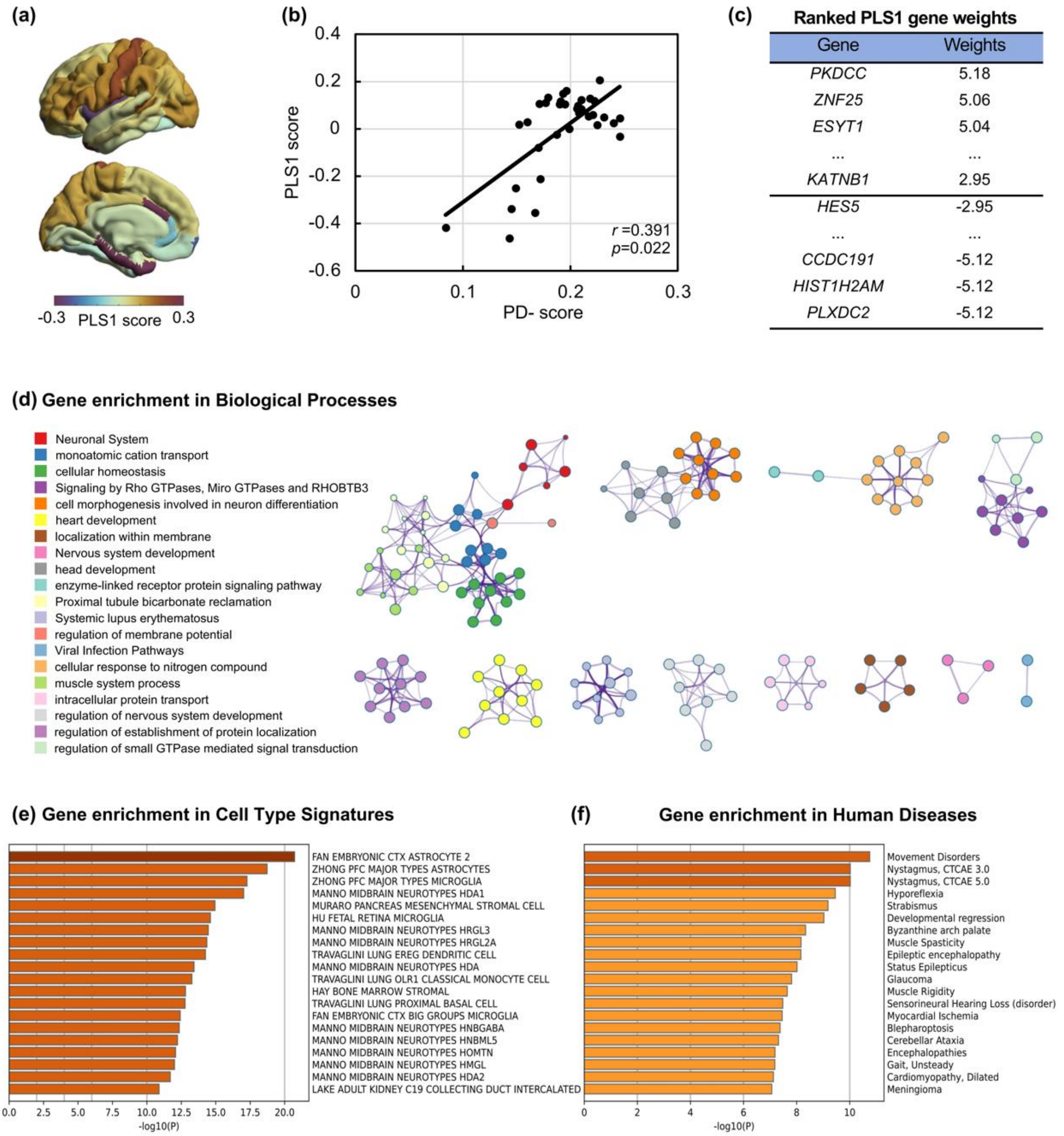


**Supplementary Figure 2**. **Partial least squares (PLS) correlation analysis between individual deviation and human brain gene expression data. (a)** Spatial map of the first component (PLS1) in the left hemisphere. **(b)** The spatial correlation between the PLS1 score and individual deviation in schizophrenia, measured by PD-score. **(c)** The ranked gene weights of PLS1 (FDR $p < 0.05$). **(d)** Top 20 gene ontology (GO) biological processes by enrichment analysis. Circle color: different biological processes. Circle size: number of genes. **(e)** Gene enrichment analysis in Cell Type Signatures. **(f)** Gene enrichment analysis in human disease-associated database.

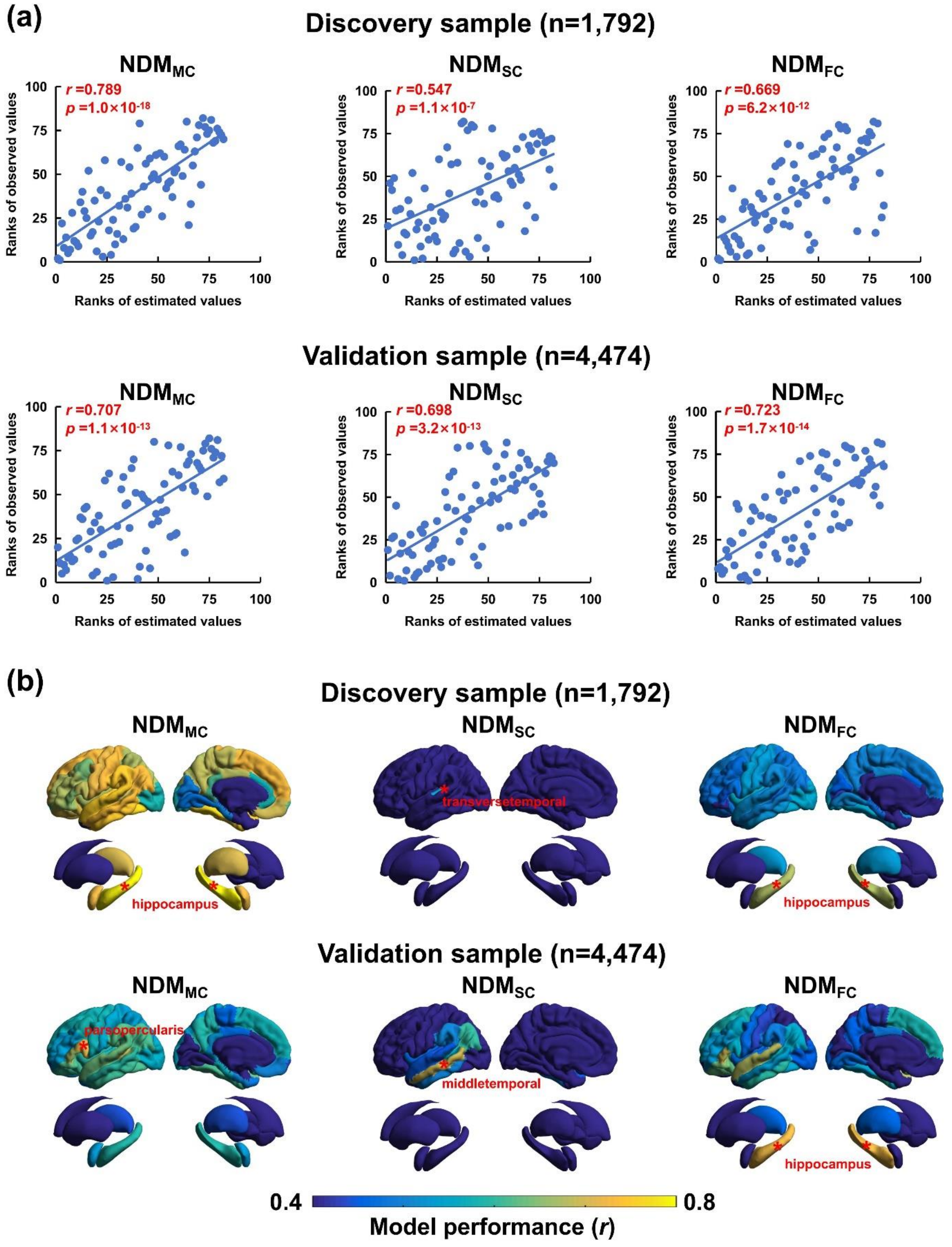


**Supplementary Figure 3**. **Performance of network diffusion model (NDM) on estimating cross-sectional GMV differences in group-level patients with schizophrenia**. **(a)** Model performance is indexed by the Spearman correlation coefficient (*r* value) between observed values and estimated values across brain regions. **(b)** Model performance with each bilateral region as the source seed of NDM. The *r* values are mapped to a brain template to show model performance of each region being the NDM seed. Red stars represent the optimal seed region (bilateral) with best performance. $NDM_{MC}$, morphological covariance connectome type-based NDM; $NDM_{SC}$, structural connectome type-based NDM; $NDM_{FC}$, functional connectome type-based NDM.
